# Oral Rinse Sourced Microbiota in Oral Health and Disease in a Representative U.S. Adult Population

**DOI:** 10.1101/2025.09.11.25335610

**Authors:** Yu Xie, Alejandro Artacho, Xiaoyu Yu, Mengning Bi, Hairui Li, Yuan Li, Andrea Roccuzzo, Alex Mira, Bob T. Rosier, Maurizio S. Tonetti

## Abstract

This study reports genus-level oral rinse microbiota profiles in a population of 3,770 U.S. adults from NHANES 2009–2012. Oral conditions explained more microbial variance than host factors, with diversity and composition differing across health, caries, periodontitis, co-occurring disease, and edentulism. In periodontitis, diversity increased alongside a shift toward anaerobic, inflammation-adapted communities. Additionally, dysbiosis-associated genera increased with disease severity gradient while health-associated ones decreased. A machine learning model based on microbial features achieved moderate accuracy in identifying severe periodontitis (AUROC in leave-one-dataset-out validation = 0.81, 95% CI 0.75-0.87; AUROC in external validation = 0.83, 95% CI 0.77-0.88). Key contributing taxa included *Desulfobulbus*, *Defluviitaleaceae* UCG-011, *Treponema*, *Filifactor*, *Pseudoramibacter*, *Mycoplasma*, *Porphyromonas*, *Bergeyella*, and *Bifidobacterium* were defined in the model. These findings support oral rinses as a non-invasive tool for monitoring oral microbial ecology and assessing the presence and severity of oral disease at the population level.

## Introduction

The oral cavity is a microbially rich environment with multiple colonization niches, including tooth surfaces, the tongue, and the oral mucosa^1^. The Human Oral Microbiome Database currently contains 770 species, whereas an individual typically harbors a much smaller subset, comprising 100-200 species (www.homd.org). Oral microbial communities exhibit strong site variability, with distinct composition profiles that are affected by local environmental conditions, including oxygen tension, pH, nutrient availability, immune system activity, and surface structure^2^. Among these niches, the supragingival and subgingival tooth surfaces are particularly exposed to microbial accumulation due to their fixed, non-shedding surfaces and complex topography. This structural stability supports the formation and maturation of biofilms, fostering relatively stable microbial communities that regrow after oral hygiene under normal conditions^3,4^.

In oral health, the oral microbiota and the host maintain a dynamic equilibrium. However, ecological imbalances can lead to a shift in the microbiota towards a pathogenic state, either locally or throughout the mouth. Dysbiosis of tooth-associated microbial biofilms initiates both caries and periodontitis^5,6^. These oral diseases rank among the most prevalent non-communicable diseases globally, impacting billions of individuals of all ages. Both arise in the oral cavity and significantly contribute to the global burden of disease, accounting for a considerable loss of disability-adjusted life years (DALYs) and are directly responsible for up to ten percent of total medical expenses^7^. Beyond oral health, these oral diseases also affect nutritional intake, quality of life, and increase the risk of systemic diseases^8–11^.

For caries, the primary disease driver is the consumption of sugars, which the oral microbiota ferments into organic acids (e.g., lactic acid) that decrease the pH and solubilize the tooth enamel^12^. By providing nutrition and changing the environment, frequent sugar intake gives a selective advantage to saccharolytic, acidogenic and aciduric bacteria that further decrease the pH, creating a vicious cycle^2^. Caries-associated bacteria include but are not limited to representatives of the genera *Streptococcus*, *Veillonella*, *Lactobacillus*, *Bifidobacterium,* and *Actinomyces*, which can contribute to pH decreases that demineralize enamel^12,13^. While in periodontal diseases, dental plaque accumulation above and below the gingival margin can trigger an inflammatory and immune response, that favors anaerobic, inflammophilic and proteolytic bacteria^2^. The dysbiosis of periodontitis is characterized by an increase in periodontitis-associated genera, like *Porphyromonas*, *Fusobacterium*, *Tannerella*, *Treponema*, *Filifactor*, and *Fretibacterium,* which become prominent in the subgingival biofilms while bacterial diversity also increases^14^.

The United States National Health and Nutrition Examination Survey (NHANES) is a nationally representative program that employs a multistage probability sampling design to gather health-related data from the non-institutionalized U.S. population^15^. Since 1999, NHANES has offered continuous cross-sectional data, including questionnaires, physical examinations, and laboratory measurements. During the consecutive cycles of 2009-2010 and 2011-2012, oral rinse samples were collected and analysed using 16S rRNA gene amplicon sequencing, producing genus-level microbial profiles for thousands of participants^16,17^.

While 16S rRNA sequencing offers limited taxonomic resolution and read length, its application in oral rinse samples from such a large population presents a unique opportunity to enhance our understanding of the oral microbiota and its connection to oral diseases and overall health. Oral rinse sampling collects microbiota from various oral sites, including the tongue, mucosa, supragingival tooth surfaces, and subgingival microbes transported into the oral cavity by the gingival crevicular fluid (GCF) ^18^. Thus, an oral rinse offers a composite snapshot of the oral, supragingival, and subgingival microbial environments. Given its non-invasive and straightforward nature, oral rinse sampling holds significant potential for population-level surveillance and microbiome-based diagnostics of oral diseases. A recent population study has shown that oral microbiota are associated with socio-economic factors and medication use^19^. However, its relationship to oral health conditions defined by clinical criteria remains underexplored.

To date, most oral microbiome studies have focused on specific intraoral sites, either caries or periodontitis, and typically within small to moderate-sized populations^20–22^, which limits the generalizability of their findings. Although these studies have enhanced the mechanistic understanding through deep sequencing of local niches, large-scale evaluations of oral microbiota patterns remain scarce. Consequently, population-level insights into microbial dysbiosis across various oral conditions are still underdeveloped.

Therefore, this study aims to profile differences in microbiota sourced from oral rinses across various oral conditions, utilizing data from the United States NHANES database. The main objective of this study is to characterize oral rinse microbiota profiles across distinct oral conditions, including oral health, dental caries alone, periodontitis alone, a combination of caries and periodontitis, and edentulism. Additionally, it aims to further assess the relationship between oral rinse-sourced microbiota and the severity of periodontitis. Lastly, the study aims to explore key periodontitis-associated taxa in the oral rinse and evaluate their potential diagnostic value for periodontitis through a machine learning approach.

## Results

### 1. Study Population and Characteristics of Subjects

A total of 9847 subjects with oral rinse-derived 16S rRNA gene amplicon sequencing data were initially considered from the NHANES 2009-2012 dataset. After applying the exclusion criteria detailed in the Method section, 6077 subjects and 1248 low-prevalence taxa were excluded. The final analytic cohort consisted of 3,770 participants with complete oral condition examination records and microbiota profiles, each with a minimum of 5,000 reads. A total of 77 genus-level taxa were retained for further analysis (**Figure S1**).

**Figure 1A** illustrates the availability of oral condition records across the dataset. **Figure 1B** shows how oral conditions, excluding edentulism, were categorized based on the presence and absence of caries and periodontitis, and how these groupings varied depending on the definition of periodontitis used. Under the Application of the 2018 Periodontal Status Classification to Epidemiological Survey data (ACES) classification system, which applies more inclusive diagnostic criteria, a larger proportion of subjects were classified as having periodontitis, resulting in a smaller proportion of individuals labeled as having oral health or caries only. In contrast, the Centers for Disease Control and Prevention-American Academy of Periodontology (CDC/AAP) classification system applied a stricter threshold, leading to a more even distribution across oral condition categories.

**Figure 1.**
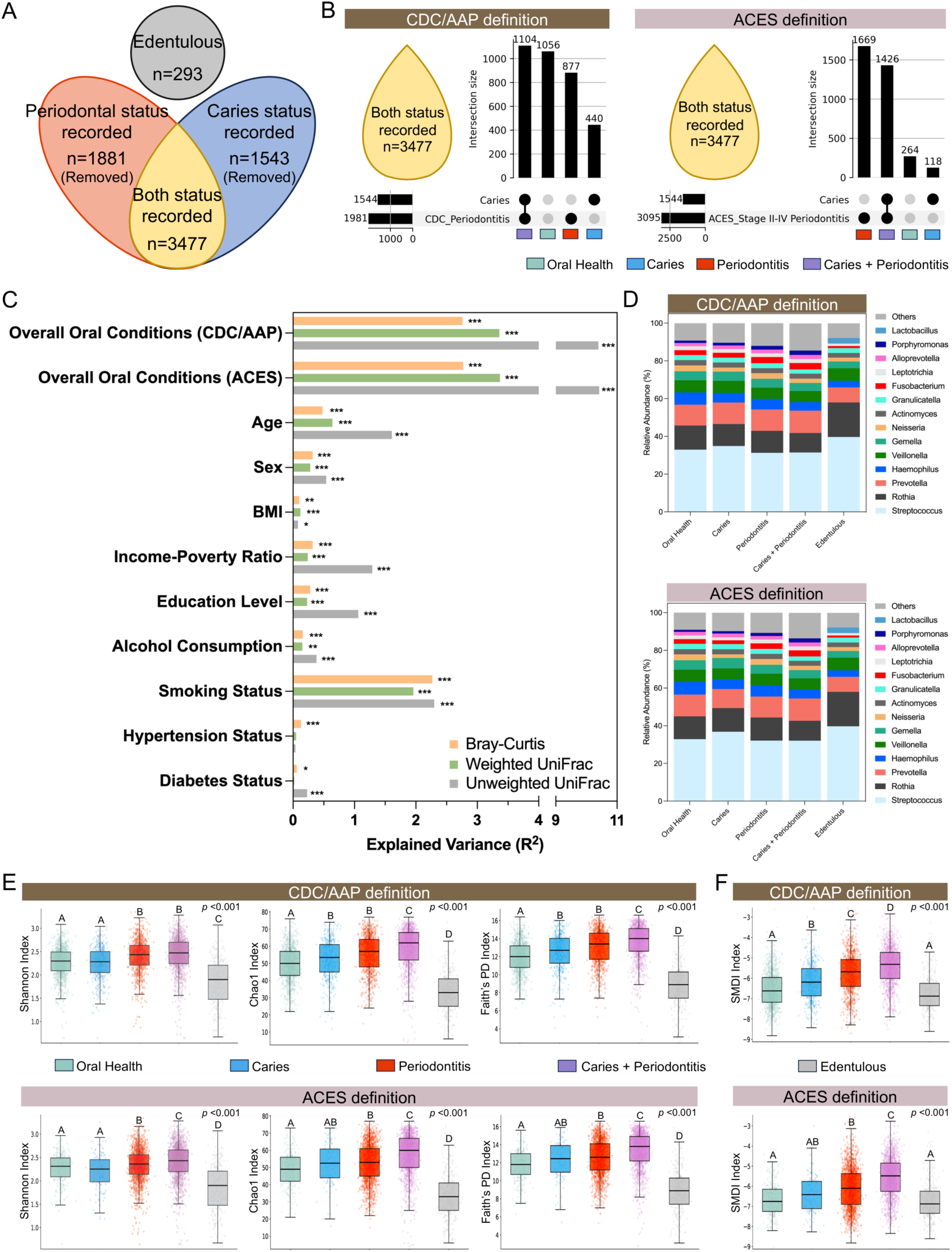
Association between oral conditions and oral microbiome. (A) Availability of oral health records based on clinical periodontal and dental examination among NHANES 2009-2011 subjects. (B) Classification of oral conditions based on caries and periodontitis status according to the CDC/AAP and ACES periodontitis definitions. (C) Variance of microbial community based on Bray-Curtis dissimilarities, weighted UniFrac distances, and unweighted UniFrac distances explained by oral health status, demographic factors, and selected general health conditions. R² and p values were calculated by PERMANOVA test. (D) Average relative abundances of the most dominant taxa (>1% average relative abundance) in different oral health statuses. (E) Alpha diversity comparisons across oral conditions, measured by Shannon, Chao1, and Faith’s PD indices. (F) Changes in periodontitis-related microbiome dysbiosis across oral conditions, measured by the subgingival microbial dysbiosis index (SMDI). In (C), significance levels are denoted as *: p <0.05, **: p <0.01, ***: p <0.001. For (E) and (F), the p-value represents the results of the Kruskal-Wallis test across all groups. Post-hoc pairwise comparisons were conducted using Dunn’s test, with different letters above bars representing statistical differences (p <0.05), while identical letters indicate no significant difference. All statistical analyses accounted for the NHANES complex survey design.

Participant characteristics stratified by different oral conditions, as defined by either the CDC/AAP or the ACES criteria, are summarized in **Tables S1 and S2**. As expected, edentulous individuals represented the oldest subgroup, with a median age of 61 years (interquartile ranges, IQR: 54-65). Under both definitions, subjects with periodontitis, either alone or in combination with caries, tended to be older than those with oral health or caries-only. The oral health group included a higher proportion of females (60.5% under the CDC/AAP definition; 68.9% under the ACES definition) and had the highest levels of education and the lowest income-to-poverty ratio. Lifestyle and systemic health characteristics also differed significantly across all oral condition groups (all *p* < 0.001), with the oral health group generally displaying lower rates of smoking, hypertension, and diabetes. Interestingly, in both demographic and systemic health profiles, individuals with caries only resembled the oral health group more closely than individuals with periodontitis (both those with periodontitis alone and those with both caries and periodontitis) under both definitions of periodontitis.

## 2. Associations Between Oral Conditions and Oral Rinse Microbiota Profiles

Although various factors can influence the oral microbial community, oral conditions proved to be the most significant explanatory factor under both the CDC/AAP and ACES definitions across Bray-Curtis, weighted UniFrac and unweighted UniFrac distances (R² = 2.76% - 10.42%, *p* < 0.001) (**Figure 1C**). Demographic factors and health conditions explained only a relatively small proportion of the variance in the oral rinse-sourced microbial community, with age (R² = 0.48% - 1.61%, *p* < 0.001), smoking (R² = 1.96% - 2.30%, *p* < 0.001) emerging as secondary explanatory factors, while body mass index (BMI), income, lifestyle, and systemic diseases contributed only minimal variation. Principal coordinates analysis (PCoA) based on Bray-Curtis dissimilarities (**Figure S2A**), weighted UniFrac distances (**Figure S2B**) and unweighted UniFrac distance (**Figure S2C**), as well as principal component analysis (PCA) based on Aitchison dissimilarities from centered log-ratio-transformed (CLR) data (**Figure S2D**), all revealed clear separation between edentulous individuals and other dentate groups. While distinctions among dentate oral condition groups remained statistically significant, though less pronounced (**Figure S2E-H**). Further analysis of microbial variation among individuals with caries, periodontitis, and comorbid caries and periodontitis showed different patterns under the two periodontitis definitions. Using the CDC/AAP definition, the caries group exhibited greater dissimilarity from the combined caries & periodontitis group than from the periodontitis group, as determined by PERMANOVA. In contrast, the ACES definition showed the opposite trend. These patterns were consistent across both relative abundance and CLR-transformed data (**Figure S3A–B**). Moreover, community-level dissimilarities between the caries and combined caries & periodontitis groups were consistently greater than those between the periodontitis and combined caries & periodontitis groups (**Figure S3C–D**).

The relative abundances of the dominant taxa (>1%) varied across different oral conditions (**Figure 1D**). While the overall composition remained relatively stable among dentate individuals, edentulous subjects showed distinct shifts. In this group, *Lactobacillus* had an average relative abundance of more than 1%, while *Porphyromonas* and *Alloprevotella* fell below 1%. Additionally, the proportions of *Streptococcus* and *Rothia* were higher in edentulous subjects. Across the remaining dentate individuals, *Streptococcus*, *Rothia*, and *Prevotella* remained the most dominant genera, regardless of the oral condition, making up over 50% of the oral rinse microbiota on average.

Microbial alpha diversity varied significantly across oral condition groups according to both CDC/AAP and ACES definitions (**Figure 1E**). Post-hoc Dunn’s tests confirmed significant pairwise differences among multiple groups. Among these, edentulous individuals displayed the lowest diversity, while those in the caries & periodontitis group exhibited the highest Shannon, Chao1 and Faith’s PD indices. Compared to individuals with oral health, those with periodontitis (either alone or in combination with caries) demonstrated significantly greater microbial richness and evenness. In contrast, the caries-only group showed minimal differences in alpha diversity compared to oral health. The subgingival microbial dysbiosis index (SMDI) further highlighted these group-level distinctions (**Figure 1F**). Individuals with periodontitis or caries & periodontitis exhibited significantly elevated dysbiosis scores under both definitions of periodontitis. In contrast, edentulous subjects had SMDI values comparable to those of the oral health group.

Given the more even group distribution under the CDC/AAP definition, along with the similar microbiome patterns observed in both classification systems, subsequent analyses concentrated on oral conditions defined by the CDC/AAP criteria in the main text, with results based on ACES presented in supplementary figures.

Taxa correlations analysis revealed shifts in intra-community microbial associations across different oral conditions (**Figure S4-S7**). Most correlations among taxa were positive, particularly at moderate to high strength (|ρ| value ≥ 0.4). Unsupervised hierarchical clustering consistently grouped taxa associated with different oral health status. Health-associated taxa (e.g. *Abiotrophia*, *Actinobacillus*, *Bergeyella*, *Capnocytophaga*, *Cardiobacterium*, *Corynebacterium*, *Haemophilus*, *Lautropia*, *Neisseria*) formed main clusters across all oral conditions. Similarly, caries-associated taxa (including *Prevotella*, *Lactobacillus*, *Veillonella*, *Atopobium*, *Actinomyces*, *Cryptobacterium*, and *Bifidobacterium*) and periodontitis-associated taxa (including *Treponema*, *Tannerella*, *Porphyromonas*, *Filifactor*, *Fusobacterium*, *Desulfobulbus*, *Mycoplasma*, *Parvimonas*, and *Eubacterium*) each formed stable clusters.

Adjusted linear regression models (**Figure 2 and Figure S8**) revealed consistent condition-specific taxonomic associations across both definitions. Compared to individuals with oral health, edentulous individuals exhibited broad negative correlations with most taxa, except for a small group that showed positive correlations, including *Streptococcus*, *Rothia*, *Bifidobacterium*, *Parascardovia,* and *Lactobacillus*. This result aligned with the low alpha diversity observed in edentulous individuals. In dentate disease groups, which included those with caries only, periodontitis only, and caries & periodontitis, most taxa exhibited positive correlations with the presence of disease. These groups also demonstrated an enrichment of anaerobic genera often linked to oral dysbiosis, such as *Fusobacterium*, *Treponema*, and *Porphyromonas*, while periodontal health-associated taxa like *Rothia* and *Actinomyces* showed negative associations. The pattern of taxonomic associations was similar between periodontitis only and caries & periodontitis, although stronger correlations were observed in the latter.

**Figure 2.**
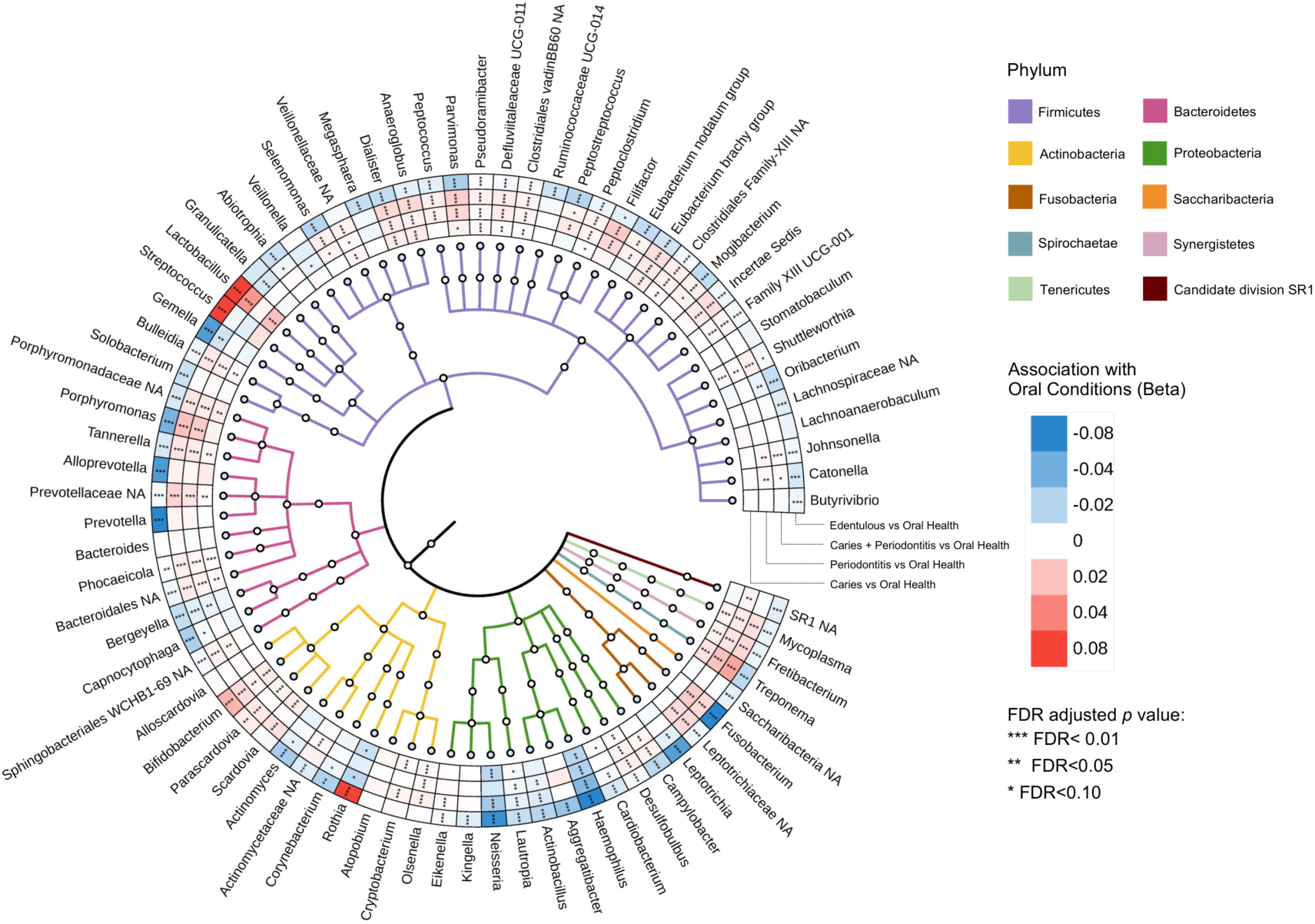
Interactive tree of life visualization of oral health-associated microbial taxa. The diagram illustrates the taxa positively associated (blue boxes in the outer wheels) and negatively associated (red boxes) in relation to oral health compared to edentulism, caries, periodontitis, or the co-occurrence of caries and periodontitis within microbial phylogenetic trees. In the heatmap, color intensity indicates the strength and direction of associations between taxa and oral conditions (each compared to oral health). Asterisks within each box indicate the significance of associations evaluated with linear regression models (adjusted for sex, body mass index, income-to-poverty ratio, education level, diabetes, and hypertension). The analysis accounted for the NHANES complex survey design. P-values have been adjusted for multiple comparisons using the false discovery rate method (FDR) with a target rate of 0.1. Periodontitis status is defined using the CDC/AAP criteria. For analyses, relative abundance data were normalized using arcsine square root (Arcsin-Sqrt) transformation. Colors on the phylogenetic tree’s clade denote different bacterial phyla.

Differential abundance analysis utilizing both DESeq2 (**Figure 3** and **Figure S9**) and ALDEx2 (**Figure S10** and **S11**) identified a substantial number of taxa with significant alterations across disease groups. As shown in the volcano plots, many taxa exhibited significant differences in abundance. For clarity, the bar plots display only those taxa with a fold change greater than 0.5 and an adjusted *p* value < 0.05 in the DESeq2 analysis. While the ALDEx2 results were summarized by presenting all taxa with an adjusted *p* value < 0.05, regardless of effect size. A complete list of differentially abundant taxa identified by DESeq2 and ALDEx2 is provided in **Supplementary Spreadsheet Tables 1–2**.

**Figure 3.**
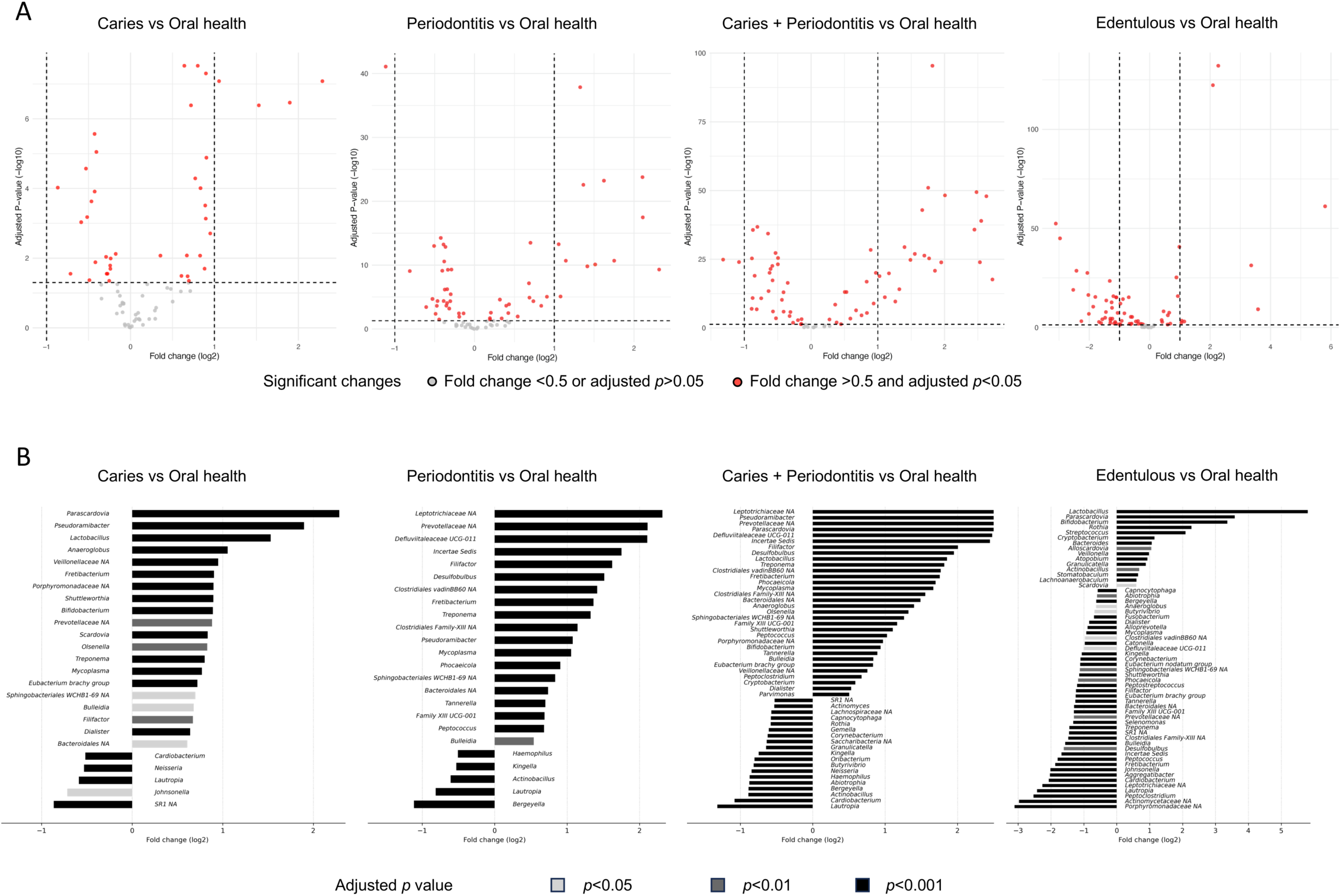
Differential abundance of taxa across oral healthy and diseased groups. (A) Volcano plots displaying differential abundance comparisons between each oral condition and oral health. Taxa with an adjusted p value < 0.05 and absolute log₂ fold change > 0.5 are highlighted in red. (B) Bar plots presenting significantly altered taxa across oral conditions, with bar colors indicating different levels of statistical significance based on adjusted p values. Comparisons were performed using DESeq2 on raw read count data, followed by false discovery rate (FDR) correction for multiple testing. Log₂ fold changes estimated by the DESeq2 model were directly used for visualization in volcano and bar plots. Oral conditions compared to oral health included caries only, periodontitis only, combined caries and periodontitis, and edentulous status. Periodontitis was defined according to the CDC/AAP case definition. Please see Figure S9 in the Appendix for the analysis using ACES to define periodontitis with DESeq2 analysis, and Figures S10 and S11 in the Appendix for differential abundance with ALDEx2 analysis.

In caries, well-known acidogenic taxa such as *Parascardovia*, *Lactobacillus*, *Bifidobacterium*, and *Scardovia,* as well as *Treponema,* were consistently more abundant, while *Neisseria* showed reduced abundance. For periodontitis, taxa enriched in disease groups included *Defluviitaleaceae* UCG-011, *Filifactor*, *Desulfobulbus*, *Treponema*, *Pseudoramibacter*, *Mycoplasma*, *Tannerella*, and *Peptococcus*, while *Bergeyella* showed consistent reduction across analyses. In addition, more significant reductions in health-associated taxa were revealed in ALDEx2 anlaysis in the diseased group compared to DESeq2 anlaysis, including *Rothia*, *Neisseria*, *Haemophilus*, *Cardiobacterium*, and *Kingella*. The combined caries & periodontitis group showed consistent enrichment of both caries- and periodontitis-associated taxa: *Pseudoramibacter*, *Parascardovia*, *Defluviitaleaceae UCG-011*, *Filifactor*, *Desulfobulbus*, *Lactobacillus*, *Treponema*, *Mycoplasma*, *Anaeroglobus*, *Olsenella*, *Bifidobacterium*, and *Tannerella*, accompanied by corresponding reductions in *Cardiobacterium*, *Bergeyella*, *Haemophilus*, *Neisseria*, and *Rothia*. In contrast to oral diseases in dentate individuals, edentulous subjects exhibited a distinct microbial shift in oral rinse. The majority of significantly altered taxa, many of which are strongly associated with caries and periodontitis, displayed decreased abundance. *Streptococcus*, *Rothia*, and only a subset of disease-related taxa such as *Lactobacillus*, *Parascardovia*, and *Bifidobacterium* remained consistently enriched, as also supported by correlation analysis (**Figure 2 and S8**). These findings highlight a markedly different microbial composition in edentulous individuals compared to those with tooth-associated disease, reflecting a transition from plaque-dominated to mucosa-associated microbiota when teeth are absent.

## 3. Oral Rinse Microbiota Reflects Periodontitis Severity

To evaluate whether oral rinse microbiota reflected the severity gradient of periodontitis, participants were stratified into different periodontitis severity categories using both CDC/AAP and ACES definitions (**Table S3 and S4**). Parallel shifts in sociodemographic and systemic health characteristics were observed with increasing periodontitis severity under both classification systems, providing epidemiological context for the corresponding microbial changes.

According to the CDC/AAP definition, alpha diversity indices (Shannon, Chao1 and Faith’s PD) exhibited a progressive increase from no periodontitis to severe periodontitis groups (**Figure 4A**). Similarly, SMDI scores rose across severity stages (**Figure 4B)**, although no statistical differences were noted between mild and moderate periodontitis in these indices. Beta diversity analyses further corroborated these findings: the PCoA plot based on Bray-Curtis dissimilarities displayed separation between periodontitis severity stages (R² = 1.30%, *p* < 0.001; **Figure 4C**), with more marked structural differences observed in the severe periodontitis group (**Figure S14A**). Similar trends were evident under the ACES definition (**Figure S12A–C, Figure S14B**). These patterns remained consistent when employing Aitchison dissimilarities calculated from CLR-transformed data (**Figure S13A**), with significant differences identified under both CDC/AAP (R² = 2.30%) and ACES (R² = 1.52%) definitions (both *p* < 0.001), and increased explained variance on more advanced periodontitis group (**Figure S14C-D**).

**Figure 4.**
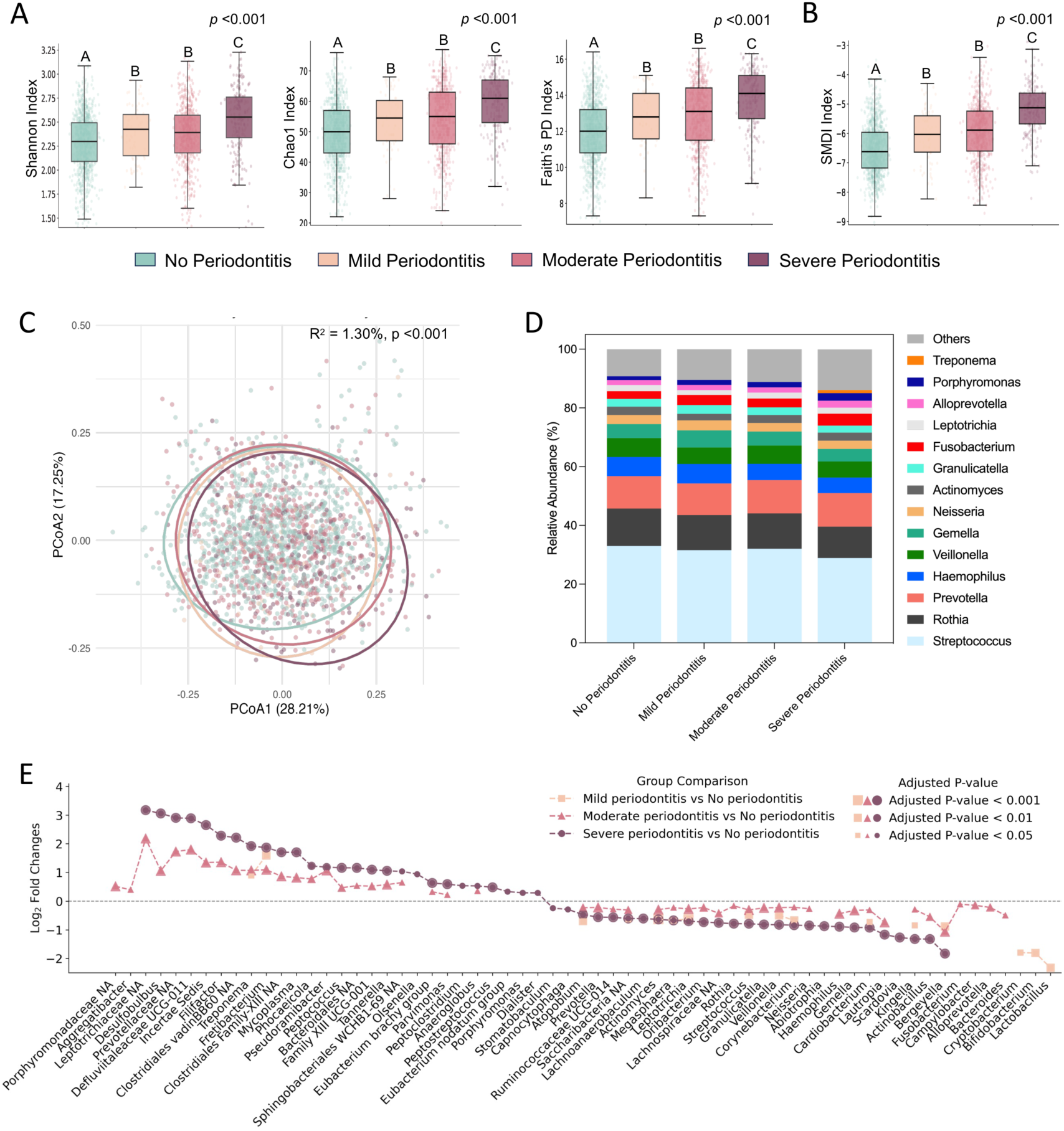
Dose-dependent association between oral microbiome profiles and periodontitis severity. (A) Alpha diversity comparisons across periodontitis severity status, measured by Shannon, Chao1 and Faith’s PD indices. (B) Changes in periodontitis-related microbiome dysbiosis across periodontitis severity status, measured by the subgingival microbial dysbiosis index (SMDI). (C) Principal coordinate analysis (PCoA) plot based on Bray-Curtis dissimilarities of relative abundance data, comparing microbial community structures across periodontitis severity status. Explained variance (R²) and p values were calculated by PERMANOVA test. (D) Average relative abundances of the most dominant taxa (>1% average relative abundance) across periodontitis severity status. (E) Differential abundance comparisons between relatively healthy periodontal status (no periodontitis in CDC/AAP definition) and more advanced periodontitis stages. Each dot represents a taxon with significant changes, with dot position indicating the magnitude of change (log₂-transformed fold change) and dot size reflecting adjusted p value significance. Comparisons of the differential abundance were performed using the DESeq2 analysis on read count data, followed by false discovery rate (FDR) correction for multiple testing. Taxa with adjusted p value < 0.05 underwent log₂ transformation for visualization. For (A) and (B), p values represent Kruskal-Wallis test results across all groups. Post-hoc pairwise comparisons were conducted using Dunn’s test; different letters above bars indicate statistically significant differences (p < 0.05), while identical letters indicate no significant difference. All statistical analyses accounted for the NHANES complex survey design.

The relative abundances of the dominant taxa (>1%) remained stable across periodontitis severity categories (**Figure 4D and Figure S12D**). *Streptococcus*, *Rothia*, and *Prevotella* consistently accounted for more than 50% of the oral rinse microbiota at all stages. However, some shifts were noted: *Treponema* emerged as a dominant taxon (>1%) in the most severe periodontitis group. Additionally, the proportion of less abundant taxa (“Others”) rose with increasing disease severity.

Differential abundance analysis among periodontitis severity groups identified specific taxa with dose-dependent behavior. Dot plots (**Figure 4E** and **Figure S12E**) illustrated the magnitude of changes through dot position and indicated statistical significance via dot size. Taking the DESeq2 analysis with the CDC/AAP definition for example, 63 taxa showed significant changes progressing from no periodontitis to moderate and severe periodontitis. Many of these taxa have been previously associated with subgingival dysbiosis in periodontitis, including *Desulfobulbus*, *Filifactor*, *Treponema*, *Fretibacterium*, *Peptococcus*, *Tannerella*, *Eubacterium brachy* group, *Parvimonas*, *Anaeroglobus*, *Porphyromonas*, *Aggregatibacter*, and *Eubacterium nodatum* group, *Dialister*.

Differential abundance analyses using ALDEx2 provided supplementary validation (**Figure S13B**). Although fewer taxa showed significant increases, key periodontitis-associated taxa remained consistently enriched along the disease severity gradient, including *Fretibacterium*, *Treponema*, *Filifactor*, *Desulfobulbus*, *Tannerella*, *Anaeroglobus*, *Peptococcus*, and *Dialister*. Conversely, a broader set of periodontal health-associated taxa showed significant decreases with increasing severity, such as *Scardovia*, *Lautropia*, *Neisseria*, *Kingella*, *Corynebacterium*, *Gemella*, *Haemophilus*, and *Rothia*. However, it was noted that these ALDEx2 analyses detected some taxa with previously known positive associations with periodontitis (e.g., *Prevotella* and *Fusobacterium*) showing significant downregulation with disease advancement. These observations suggest that results based on CLR-transformed data (ALDEx2), while broadly consistent with findings from DESeq2 analysis, should be interpreted cautiously.

## 4. Diagnostic Potential of Periodontitis-Associated Taxa Identified by Machine Learning

To further investigate the taxa significantly associated with periodontitis severity and to evaluate the diagnostic potential of oral rinse microbiota, we developed machine learning model using random forest classifiers. Feature selection was performed with least absolute shrinkage and selection operator (LASSO) regression on the training dataset, which comprised 70% of the samples stratified by disease severity (ratio 5:3:1). Model performance was validated both internally and externally: internally using a leave-one-dataset-out (LODO) testing set consisting of the remaining 30% of samples with the same severity ratio, and externally using a local cohort established by the researchers (with a severity ratio of approximately 1:5:1) (**Figure 5A**). The random forest models were trained and validated using both CLR-transformed data and on relative abundance data, to evaluate how data type influences predictive performance and interpretability across internal and external validation.

**Figure 5.**
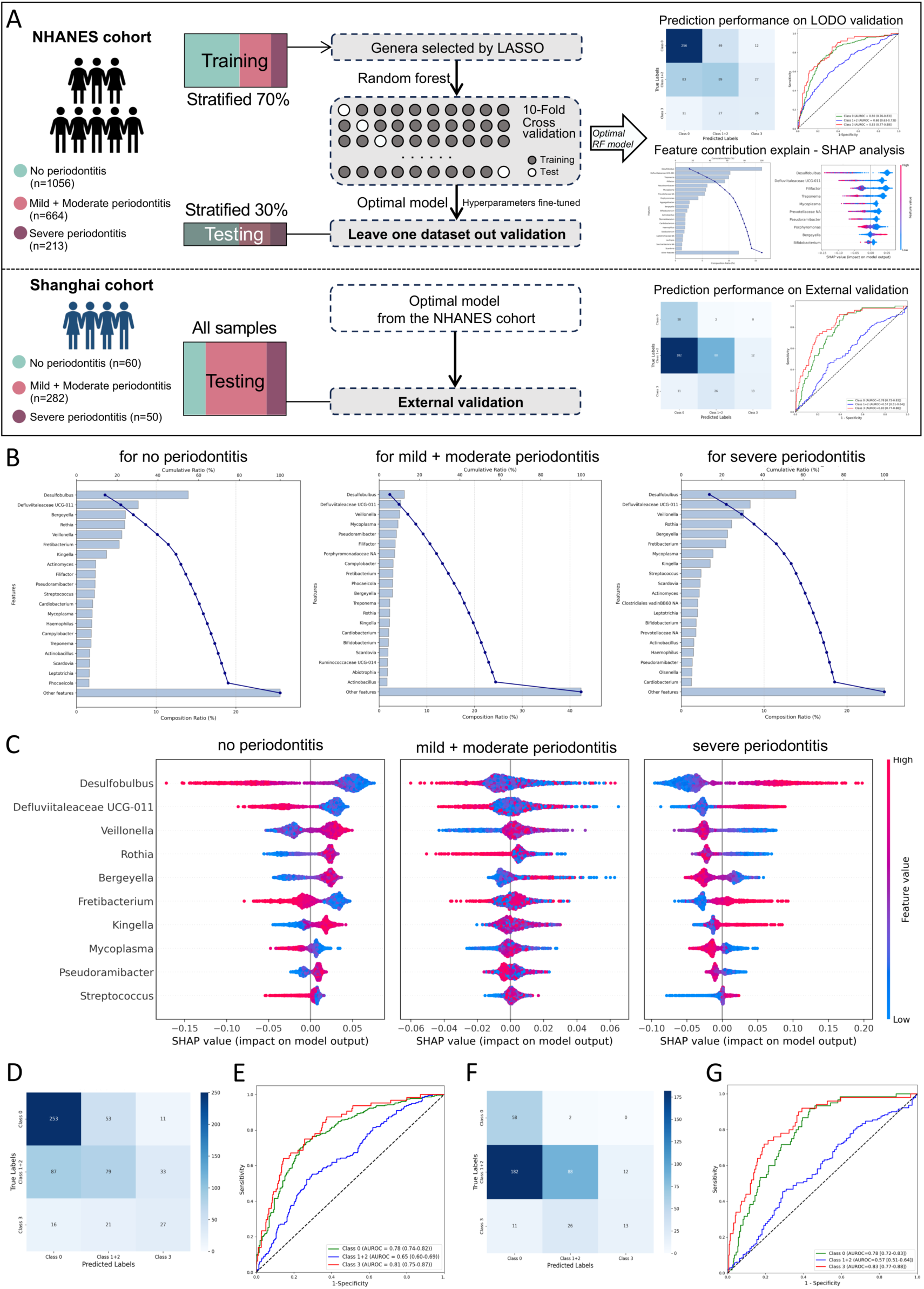
Machine learning model for periodontitis prediction based on microbial taxa. (A) Framework for data partitioning, model training, independent leave-one-dataset-out (LODO) validation, and external validation, utilizing genus-level features selected by LASSO regression for model construction. Illustrated using different periodontitis severity groups defined by the CDC/AAP definition on CLR-transformed data for an example. (B) Pareto plots showing the single and cumulative contribution of dominant taxa to model interpretability, as determined by the random forest multi-class model. (C) Shapley additive explanation (SHAP) summary plots for the top 10 taxa contributing to model interpretability. Each point represents a sample, colored by the CLR transformed relative abundance of the taxon (blue to red representing low to high abundance). The x-axis shows the SHAP value, indicating the magnitude and direction of each taxon’s impact on the model output. (D) Confusion matrix displaying the performance of the trained random forest multi-class model in LODO validation. (E) Receiver operating characteristic (ROC) curves of the trained random forest multi-class model for distinguishing periodontitis severity categories in the LODO validation. (F) Confusion matrix displaying the performance of the trained random forest multi-class model in external validation. (G) ROC curves of the trained random forest multi-class model for distinguishing periodontitis severity categories in external validation.

In LODO validation, the optimized models achieved acceptable classification performance for differentiating severity stages of periodontitis (**Figure 5D-E, Figure S15**). Under the CDC/AAP definition, the area under receiver operating characteristic curve (AUROC) for classifying the least severe group (no periodontitis) was 0.78 (95% CI: 0.74–0.82), and 0.81 (95% CI: 0.75–0.87) for the most severe group (severe periodontitis). Under the ACES definition, AUROCs were slightly lower: 0.73 (95% CI: 0.68–0.79) for Stage I periodontitis and 0.73 (95% CI: 0.68–0.77) for Stage III–IV periodontitis. However, AUROC values for intermediate severity categories were lower in both systems: 0.65 (95% CI: 0.60–0.69) for mild and moderate periodontitis (CDC/AAP) and 0.59 (95% CI: 0.54–0.63) for Stage II periodontitis (ACES). Across all comparisons, diagnostic performance was consistently slightly higher with the CDC/AAP definition. Models based on relative abundance data showed similar performance in internal validation (**Figure S15**).

In external validation, the optimal model derived from the NHANES dataset demonstrated acceptable performance on CLR-transformed data (**Figure 5F-G, Figure S15A**). For the CLR-based model, the local cohort showed similar or even improved performance in the least severe group (AUROC in no periodontitis under CDC/AAP definition = 0.78 [0.72-0.83]; AUROC in stage I periodontitis under ACES definition = 0.76 [0.65-0.86]) and the most severe group (AUROC in severe periodontitis under CDC/AAP definition = 0.83 [0.77-0.88]; AUROC in stage III/IV periodontitis under ACES definition = 0.83 [0.70-0.92]). In contrast, performance in the intermediate groups declined modestly under both definitions (AUROC in mild+moderate periodontitis = 0.57 [0.51-0.64]; AUROC in stage II periodontitis = 0.50 [0.37-0.63]) (**Figure S15A**). By comparison, the model trained on relative abundance data consistently showed reduced performance across all severity groups in external validation (Figure S15B).

Feature importance was evaluated using Shapley Additive exPlanations (SHAP) analysis. Across both data types (CLR or relative abundance) and periodontitis definitions (CDC/AAP and ACES), the top 10 contributing taxa consistently accounted for ∼50-70% of the total feature contribution in the least and most severe categories, but only ∼40-50% in the intermediate category (**Figure 5B, Figure S16A and Figure S17A**).

Similar to patterns observed in differential abundance analyses (**Figure S10 and Figure S11)**, models trained on CLR-transformed data (**Figure 5B-C and Figure S16**) highlighted a broader set of periodontal health-associated taxa as dominant contributors to classification. Under the CDC/AAP definition, leading features included *Desulfobulbus*, *Defluviitaleaceae* UCG-011, *Veillonella*, *Rothia*, *Bergeyella*, *Fretibacterium*, *Kingella*, *Mycoplasma*, *Pseudoramibacter*, and *Streptococcus*. Under the ACES definition, major contributors comprised *Desulfobulbus*, *Filifactor*, *Phocaeicola*, *Bergeyella*, *Haemophilus*, *Rothia*, *Defluviitaleaceae* UCG-011, *Butyrivibrio*, *Fretibacterium*, and *Clostridiales vadinBB60 NA*. By contrast, models based on relative abundance data emphasized a narrower set of taxa that were predominantly associated with periodontitis, particularly in the least and most severe categories. These included *Desulfobulbus*, *Defluviitaleaceae* UCG-011, *Treponema*, *Filifactor*, *Prevotellaceae NA*, and *Porphyromonas*, with shifts in dominant contributors between the CDC/AAP and ACES definitions (**Figure S17**).

Despite these differences in the identity of top-ranked features, the directionality of impact for several key taxa was consistent across data transformations and disease definitions, such as *Desulfobulbus*, *Defluviitaleaceae* UCG-011, and *Filifactor*, particularly in the least and most severe periodontitis categories (**Figure 5C, Figure S16B, and Figure S17B**). For example, in the CLR-based diagnostic models under the CDC/AAP definition (**Figure 5C**), five of the top 10 taxa including *Desulfobulbus*, *Defluviitaleaceae* UCG-011, Fretibacterium, Mycoplasma and Streptococcus showed strong contributions toward severe periodontitis, while exhibiting an opposite trend in the “no periodontitis” group. Importantly, several of these taxa (*Desulfobulbus*, *Defluviitaleaceae* UCG-011, *Fretibacterium* and *Mycoplasma*) have been previously reported as associated with advanced disease, lending biological plausibility to their predictive rold. By contrast, contributions in the “mild + moderate periodontitis” group were less consistent. Taxa such as *Fretibacterium*, *Mycoplasma*, *Pseudoramibacter*, and *Kingella* displayed opposite or ambiguous patterns to the model prediction, underscoring the difficulty of reliably predicting intermediate disease states.

## Discussion

This study provides a comprehensive analysis of oral rinse-derived microbiota across varying oral health conditions of periodontitis, dental caries, and edentulism, within a representative U.S population. Oral rinse samples, encompassing microbial communities from diverse oral niches, offer a composite view on oral microbiota dynamics. Although previous studies by Qi et al. ^23^ and Chaturvedi et al. ^19^ using the NHANES database demonstrated that oral rinse-derived microbiota are associated with host factors such as smoking, age, sex, and socioeconomic status, our findings indicate that oral diseases exert a stronger influence on microbial community variance than these host factors. This underscores the importance of anlayzing microbial changes and their associations with different oral conditions in oral rinse samples. To our knowledge, this is the first large-scale study to comprehensively evaluate how the bacterial composition in oral rinse is affected by various oral conditions.

Among oral diseases, periodontitis was associated with increased alpha diversity and a higher SMDI, alongside specific disease-associated taxonomic shifts in oral rinse samples. Dental caries was linked to elevated Chao1 and Faith’s PD indives and distinct microbial alterations that differed markedly from those observed in periodontitis. In individuals with both periodontitis and caries, the oral rinse microbiota more closely resembled that of periodontitis, while correlation analysis highlighted distinct caries-associated and periodontitis-associated taxonomic clusters. Edentulism, characterized by the absence of teeth, was characterized by reduced alpha diversity and the decrease tooth-associated genera. Collectively, these findings suggest that oral rinse samples are effective for detecting microbial biomarkers across various oral health states. Furthermore, we showed that microbiota information derived from oral rinse carries diagnostic potential for differentiating periodontitis severity through machine learning models.

As noted in the Methods, this study applied two periodontitis classification systems to define oral conditions. Both the CDC/AAP and ACES definitions captured shifts in disease-associated microbiota. However, due to the stricter criteria of the ACES definition^24^, particularly for periodontal health and early-stage disease, no individuals in the NHANES 2009-2012 dataset met the “no periodontitis” criteria. As a compromise, 382 individuals with stage I periodontitis were designated as “periodontal health”, which resulted in an imbalanced group distribution under the ACES framwork. This observation is consistent with findings by Tay et al., who reported that the ACES system may be overly stringent for defining periodontal health and may overestimate periodontitis prevalence, as in their study for NHANES 2009-2014 cycles there are 93.1% adults defined as periodontitis under ACES framework^25^. Moreover, microbial profiles between stage I and stage II periodontitis showed limited distinction, indicating reduced discriminatory power at these early stages. Accordingly, the CDC/AAP definition was adopted for primary analyses, with ACES-based results provided in the Supplementary information.

Notably, while periodontitis is well known for its association with plaque accumulation and changes in the microbiota of subgingival plaque, we found that the oral rinse-sourced microbiota profile for periodontitis still exhibited similar patterns to those observed in subgingival plaque. The alpha diversity was significantly elevated across all periodontitis classification systems and increased with disease severity. The beta diversity also differed between individuals with different severities of periodontitis. Further analysis confirmed that oral rinse microbiota captured changes associated with periodontal diseases. The SMDI, developed initially using subgingival plaque data^26^ and later validated in saliva and tongue samples^27^, increased significantly with the advancement of periodontitis.

Taxa correlation analysis revealed distinct clustering patterns indicative of microbial network restructuring across oral conditions. Unsupervised hierarchical clustering consistently identified three major clusters corresponding to health, caries, and periodontitis. The correlations among taxa were mainly concentrated within the same health- or disease-associated clusters, rather than spanning across them. This pattern suggests that oral microbial interactions are largely shaped by shared ecological or pathological contexts, reinforcing the tendency of taxa to co-vary within similar health states. Notably, *Prevotella* consistently appeared in caries-associated clusters rather than periodontitis-associated clusters, possibly due to the context-dependent role of this genus and the sample type used in this study. Inter-taxa correlations further revealed shifts in functional network structure. In the health group, significant correlations were primarily within clusters, whereas disease groups exhibited more significant inter-cluster correlations. Health-associated taxa such as *Neisseria* often showed negative correlations with disease-related taxa, supporting their reported anti-inflammatory and colonization resistance roles^22,28^. An intriguing example is the positive correlation between *Porphyromonas* and *Neisseria* in healthy individuals, absent in periodontitis, likely reflecting the coexistence of both commensal (e.g., *P. catoniae*) and pathogenic (e.g., *P. gingivalis*) species within the genus. These findings emphasize that microbial dysbiosis in oral disease involves not only changes in taxon abundance, but also the reorganization of microbial interaction networks.

Moreover, differential abundance analysis revealed substantial overlap between our findings and taxa previously reported to be enriched in periodontitis by Feres et al. ^14^ and Meuric et al. ^29^, including *Treponema*, *Tannerella*, *Porphyromonas*, *Filifactor*, *Fusobacterium*, *Desulfobulbus*, *Mycoplasma*, *Selenomonas*, *Peptostreptococcus*, *Pseudoramibacter*, *Parvimonas*, *Eubacterium* and *Campylobacter*. While health-associated genera such as *Rothia*, *Neisseria*, *Veillonella*, *Streptococcus*, *Haemophilus,* and *Bergeyella* were reduced, in line with established cognitions. These results supported the utility of oral rinse in capturing subgingival microbiota changes. Previous studies have also shown that salivary microbiota partially reflects subgingival alterations due to the presence of gingival crevicular fluid^30,31^. However, future studies should compare the effectiveness of saliva and oral rinse microbiota profiles in discriminating between health and disease.

Building on these findings, we explored the diagnostic utility of oral rinse for periodontitis. We focused on periodontitis rather than caries due to both clinical and research considerations. Importantly, periodontitis diagnosis typically requires full-mouth periodontal probing, which is invasive, technique-sensitive, and time-consuming. Consequently, non-invasive alternatives have been widely explored, often focusing on inflammatory biomarkers (e.g., IL-1β, IL-6, MMP-8) in oral fluids^32,33^. However, these markers show limited diagnostic performance, likely due to their inability to capture the complexity of periodontal disease. Thus, given that oral rinse microbiota partially mirrors subgingival microbial patterns, we hypothesized that different stages of periodontitis can be identified based on the bacterial composition of oral rinse samples. We developed a multi-class classification model to distinguish between no periodontitis, mild + moderate periodontitis, and severe periodontitis. Although the model was based on genus-level data, it achieved moderate diagnostic performance according to the diagnostic metric AUROC. Notably, while models using CLR transformed data and relative abundance data performed similarly in internal validation, the model built on CLR data exhibited superior performance in external validation, maintaining preditive accuracy in a heterogeneous dataset. This observation aligns with findings by Li et al. in gut microbiome-based digestive disease prediction^34^, highlighting the greater robustness and generalizability of CLR-transfromed data for predictive modelling across diverse cohorts. Meanwhile, the model showed poor to fair performance in predicting intermediate disease status, which poses a challenge for its clinical application. This drawback is expected, as intermediate periodontitis represents a transitional stage of disease progressing without a distinct microbial signature for reliable detection. Nevertheless, we anticipate that increasing sequencing depth and applying machine learning at the species or even strain level could substantially improve the diagnostic performance, potentially overcoming this limitation. From another perspective, SHAP analysis highlighted the top contributing taxa, including well-known periodontitis-associated genera: *Treponema*, *Filifactor*, *Desulfobulbus*, *Porphyromonas*, *Pseudoramibacter*, and *Mycoplasma*; and the health-associated genus *Bergeyella*^14,29^, demonstrating the interpretability of the machine learning model. Although preliminary, these findings suggest the potential value of future studies employing greater sequencing depth of oral rinse samples for diagnostic purposes. Moreover, the interpretability of the model supports both the biological relevance of oral rinse microbiota and the reliability of its prediction.

In case of caries, the accumulation of acidogenic and aciduric bacteria leads to a shift in supragingival plaque toward a pathogenic biofilm, often accompanied by reduced microbial diversity due to acidic stress^5,35^. However, in oral rinse samples of this study, alpha diversity appeared similar or even elevated in individuals with caries. This may reflect the composite nature of oral rinse, which collects microbes from both diseased and healthy oral sites. The inclusion of caries-associated taxa likely contributed to the slight increase in diversity. Additionally, elevated SMDI in caries individuals may suggest subclinical periodontal inflammation, gingivitis or an increase in proteolytic bacteria, such as those involved in dentin caries^36^. Differential abundance analysis revealed an enrichment or recognized caries-associated taxa, including *Lactobacillus*, *Bifidobacterium*, *Pseudoramibacter*, *Dialister*, and *Scardovia*, and a decline in health-associated taxa sucha as *Rothia*, *Neisseria*, *Haemophilus* and *Capnocytophaga*^5,37^. In contrast, prior studies using saliva samples in a smaller population found fewer significantly different taxa between caries-active and healthy individuals^38^, indicating the potential advantage of oral rinse in capturing broader microbial signals. However, this should be validated in a large cohort through the analysis of both saliva and oral rinse samples.

Microbial profiles in individuals with both caries and periodontitis revealed complex patterns of dysbiosis. Beta diversity analysis indicated clear community divergence. A distinct dysbiotic profile emerged, characterized by the concurrent enrichment of periodontitis-associated bacteria (*Filifactor*, *Treponema*, *Fretibacterium*, *Tannerella,* and *Porphyromonas*) and caries-associated genera (*Lactobacillus*, *Bifidobacterium*, *Parascardovia*, and *Scardovia*), alongside a consistent reduction of health-associated taxa such as *Haemophilus*, *Rothia*, *Neisseria,* and *Bergeyella*. Interestingly, microbial dissimilarity analyses indicated that the caries & periodontitis group was more similar to the periodontitis group than the caries group. This pattern was echoed in taxa correlation and hierarchical clustering analyses. Taxa in the caries & periodontitis group were divided into three distinct clusters: health-, caries-, and periodontitis-related cluster. With the periodontitis-related cluster dominating, and health-associated cluster representing only a small portion of taxa. These findings suggest that oral rinse can capture overlapping microbial signatures of dual pathologies while the presence of one disease does not obscure the microbial indicators of the other. And it may more effectively reflect periodontitis status, even though key periodontitis-associated taxa mostly exist in subgingival plaque.

In edentulous individuals, as expected, significant shifts in microbiota were observed. Tooth loss removed stable hard surfaces for biofilm formation, leading to a restructured microbial habitat favoring colonization of mucosa, dentures, and the tongue dorsum. This restructuring resulted in reduced alpha diversity and a distinct microbial composition, consistent with previous findings in saliva^39^. Segata et al. ^40^ demonstrated that microbial communities differ substantially across oral niches, including sub- and supra-gingival plaque, mucosa sites (buccal mucosa, keratinized gingiva, and hard palate), and saliva, tongue, tonsils, and throat, which further supports these observations. In our analysis, most taxa decreased in the edentulous individuals, while *Lactobacillus*, *Bifidobacterium*, *Scardovia*, *Parascardovia*, *Rothia* and *Streptococcus* increased. Notably, aciduric genera associated with caries persisted, likely due to their ability to adhere to mucosal and denture surfaces and thrive in acidic environments. These shifts highlight the adaptability of oral microbiota to anatomical changes.

Several limitations should be acknowledged. First, the NHANES dataset was limited by taxonomic resolution, as it only provided genus-level information, which restricted insights into species- and strain-level associations. This also constrained the diagnostic power of our machine learning models, as genera may include both disease- and health-associated species. For example, *Leptotrichia* and *Prevotella* are known to include both periodontal health- and periodontitis-associated species^41^, while *Steptococcus* has representatives associated with caries (e.g., *S. mutans*) and others with caries-free individuals (e.g., *S. dentisani*) ^42^. Nevertheless, on a genus level, *Prevotella* is associated with periodontitis, while *Steptococcus* is associated with caries, indicating that a larger proportion of representatives within these genera are associated with the specific disease. Additionally, at the genus level, some taxa associated with caries are also linked to periodontal health, including *Streptococcus* and *Veillonella*. These observations limit the information that can be extracted from genus-level analyses, as well as the diagnostic and predictive potential. Future studies employing high-resolution sequencing (e.g. PacBio or shotgun metagenomics) are needed to capture more accurate species-level microbial profiles. Secondly, the NHANES examination data offered limited detail on clinical parameters, lacking information on caries severity, bleeding on probing during periodontal examination, and denture use in edentulous individuals, which reduced the granularity of disease classification and limited sensitivity analyses on specific subsets. Thirdly, the database only included individuals aged 18 to 69 for the 16SrRNA sequencing, limiting generalizability to older populations. In addition, the database provided only the taxonomically assigned read count table without ASVs data or batch information, restricting the possibility of updated analyses using newer reference databases or applying batch-correction methods.

Despite these limitations, our findings demonstrate that oral rinse, while less spatially precise than site-specific samples (e.g. subgingival plaque for periodontitis, supragingival plaque for caries), can be used to detect key microbial signatures of oral diseases. This is especially valuable in large-scale or population-based studies, where non-invasive, easily collected biospecimens are essential. Our random forest model showed moderate diagnostic performance in classifying periodontitis severity based on genus-level data, supporting the feasibility of oral rinse as a diagnostic tool. Future research integrating species-level data and advanced modeling may further refine its utility in clinical and epidemiological setting.

## Methods

### Study population

The US NHANES has been designed to represent the employed United States population of 164 million adults. For this study, participants from two consecutive NHANES cycles (2009-2010 and 2011-2012) who had 16S rRNA gene amplicon sequencing data from oral rinse samples were included^16,17^. Individuals were excluded if they were under 18 years old, pregnant, lacked a dental examination, did not have both caries and periodontal records, or had only one remaining tooth, which did not allow for periodontal assessment (**Figure S1**) ^24,43^. The NHANES comprises two primary components^16,17^: a household structured interview was conducted by trained personnel to gather information on demographics, dietary intake, tobacco use, and medical history; and assessments were conducted by calibrated examiners, including evaluations of body measurements, oral health status and biological sample collection at a mobile examination center (MEC). All collected biospecimens were analyzed in certified laboratories. The National Center for Health Statistics’ Ethics Review Board approved the NHANES.

The local database served as an external validation dataset for the machine learning model. This cross-sectional cohort was established by the researchers in Shanghai, China, which recruited participants between July 2023 and July 2024. Oral rinse samples and standardized periodontal examinations were collected, and a total of 392 participants were included in the external validation.

### Oral health status definitions

Participants were classified based on three components obtained through clinical examination at the MECs: dentition status, caries status, and periodontal health status. The dentition status was defined based on tooth presence; participants were classified as edentulous if all teeth were recorded as missing, while all others were considered dentate. Among dentate individuals, a caries subject was defined by the presence of at least one untreated decayed tooth, assessed as the presence of a break in the enamel surface detected by a dental explorer in any tooth or a shadow (lack of translucency) upon transillumination in the anterior teeth in the 2011-2012 cycle. In the 2009-2010 cycle, the presence of caries was defined based on the examiner’s referral for additional care.

Periodontal status was assessed through pocket depth and clinical attachment loss in the full-mouth periodontal probing at six sites per tooth. Two standard, well-established periodontal classification systems were used separately in this study: the Centers for Disease Control and Prevention (CDC)-American Academy of Periodontology (AAP) classification system and the Application of the 2018 Periodontal Status Classification to Epidemiological Survey data (ACES) system. For the CDC/AAP classification system, subjects were classified as having no periodontitis, mild periodontitis, moderate periodontitis, or severe periodontitis^43^. For ACES, participants were grouped into four categories: no periodontitis, stage I, stage II, and stage III/IV periodontitis^24^. Additional details are provided in the supplementary information.

Dentate individuals were further grouped based on the presence or absence of caries and periodontitis: (1) oral health (neither caries nor periodontitis present), (2) caries only (only caries present, without periodontitis), (3) periodontitis only (only periodontitis present, without caries), and (4) caries & periodontitis (both caries and periodontitis present). Notably, under the ACES classification system, no participants met the criteria for “no periodontitis”. Therefore, Stage I periodontitis under the ACES definition was considered representative of periodontal health for the classification of “oral health” and “caries only”.

### 16S rRNA gene amplicon sequencing

In NHANES, oral rinse samples were collected at the MECs before the oral health examination. Trained dental examiners instructed participants to rinse and gargle with 10 mL of saline mouthwash for 30 seconds. Sample DNA extraction and 16S rRNA gene amplicon sequencing were performed in certified laboratories as previously described^44^. Briefly, the V4 region of the 16S rRNA gene was amplified using primers 515F (5’-GTG CCA GCM GCC GCG GTA A-3’) and 806R (5’-GGA CTA CHV GGG TWT CTA AT-3’), producing ∼390 bp amplicons. Sequencing was conducted on the Illumina HiSeq 2500 System using 2×125 bp paired-end reads, following the manufacturer’s protocols. A total of 19 individual sequencing runs were carried out.

Raw sequencing reads were demultiplexed using QIIME1 (version 1.9.1) to generate separate forward and reverse FASTQ files for each individual. Due to insufficient overlap between the 125 bp paired-end reads, only forward reads were retained for further analysis. Sequence processing was performed using the DADA2 pipeline (version 1.2.1), which included quality filtering, error correction, chimera removal, and inference of amplicon sequence variants (ASVs). Each sequencing run was processed independently using DADA2 and ASV tables, and the results were subsequently merged. The taxonomic assignment of ASVs was carried out using the SILVA v123 database. Relative abundance and read count tables were generated with QIIME 1.9.1 without rarefying the data.

In the local cohort, oral rinse samples were collected by calibrated examiners before periodontal examination. Participants were instructed to rinse and gargle with 5 mL of distilled water for 30 seconds. DNA was extracted from the samples, and 16S rRNA gene amplicon sequencing was performed following standard protocols []. The V3-V4 hypervariable region of the 16S rRNA gene was amplified using primers 341F (5’-CCT AYG GGR BGC ASC AG-3’) and 806R (5’-GGA CTA CNN GGG TAT CTA AT-3’), generating ∼460 bp amplicons. Sequencing was conducted on the Illumina NovaSeq 6000 platform with 2×250 bp paired-end reads, according to the manufacturer’s instructions. Raw sequencing reads were processed using QIIME2 (version 2025.4.0), including demultiplexed, quality filtering, and denoising with the DADA2 pipeline to generate ASVs. Taxonomic assignment of ASVs was performed with SILVA database. Feature tables were collapsed to genus level in QIIME2, and read count tables were generated without rarefaction.

### Data analysis

Descriptive statistics analyses of clinical parameters across groups were performed using SPSS statistics (version 26.0, IBM Corp., U.S). The normality of continuous variables was assessed, and non-normally distributed data are presented as medians with interquartile ranges (IQRs) and compared using the Kruskal-Wallis test. Categorical variables are summarized as count (frequency) and compared using the Chi-square test.

For microbiome analysis, taxa were excluded if they were not present with >10 reads in ≥30% of individuals in at least one of the oral conditions (oral health, caries, periodontitis, caries & periodontitis, or edentulism), or lacking taxonomic annotation at the class level. In addition, samples with <5000 reads were removed. All microbiome analyses and visualizations were conducted using the filtered dataset (**Figure S1**).

In this study, statistical analyses and visualizations were performed using R (v4.4.1) ^45^ and Python (v3.12) ^46^. Alpha diversity indices (Shannon and Chao1) were determined using the R Vegan library^47^ and the subgingival microbial dysbiosis index (SMDI) was calculated using the online SMDI platform^26&^, both using the read count tables. Intergroup comparisons were performed using the Kruskal-Wallis test, followed by pairwise post-hoc analysis using Dunn’s test with Bonferroni correction. For beta diversity analysis, both untransformed relative abundance data and centered log-ratio (CLR) transformed data were used. Bray-Curtis dissimilarities, weighted UniFrac distances and unweighted UniFrac distances were calculated from the relative abundance data, whereas Aitchison distances were computed from the CLR-transformed data. Statistical differences in beta diversity were assessed using permutational multivariate analysis of variance (PERMANOVA). Bray–Curtis, weighted UniFrac, and unweighted UniFrac results were visualized with principal coordinates analysis (PCoA), while Aitchison distances were visualized with principal component analysis (PCA).Correlations between taxa under different oral conditions were analyzed using Spearman correlation with hierarchical clustering, corrected by false discovery rate (FDR). Associations between microbial taxa and oral health conditions were evaluated using linear regression models, with the “oral health” group serving as the reference. All models were adjusted for sex, body mass index (BMI), income-to-poverty ratio, education level, hypertension, and diabetes. Relative abundance data were transformed to the arcsine square root (arcsin-Sqrt) before the analysis. *P* values were adjusted using the Benjamini-Hochberg false discovery rate (FDR), with a significance threshold set at FDR < 0.1. The Interactive Tree of Life (iTOL, version 7.1) was used to visualize the associations combined with a phylogenetic tree. Differential abundance analyses were conducted using the DESeq2 and ALDEx2 packages in R, both applied to read count data. Comparisons of taxa between the oral health group and each disease group were performed, and p-values were adjusted for multiple testing using the Benjamini–Hochberg procedure. Analyses for diversities, SMDI, taxa-taxa correlation and taxa–oral condition associations all incorporated the complex sampling design of NHANES by applying cycle-specific examination weights (adjusted for nonresponse), along with strata and multistage-clustered sampling design, using survey package in R. As two NHANES cycles were used in this study, the examination weightes were divided by 2.

Using either CLR-transformed or relative abundance data, machine learning models were constructed using Python (version 3.12) to classify the severity of periodontitis based on oral rinse-sourced microbiota profiles. Due to the class imbalance of periodontitis severity under the CDC/AAP definition, individuals with mild and moderate periodontitis were merged into a single “mild and moderate periodontitis” group. The dataset was randomly stratified by disease severity and split into training (70%) and testing (30%)subsets. The training set was used for model development with 10-fold cross validation for hyperparameter tuning, while the testing set was reserved for leave-one-dataset-out validation. To reduce dimensionality and mitgate overfitting, least absolute shrinkage and selection operator (LASSO) regression was applied to the training set. Predictors were standardized with z-score normalization, and features with non-zero coefficients were selected for downstream modeling. The random forest (RF) classifier was trained with class-weighted learning to address imbalance. Hyperparameters were optimized using GridSearchCV with cross-validation, targeting the optimal weighted F1 score. For external validation, the optimal RF model was applied to the local cohort dataset. Taxa included in the model but absent from the local cohort were treated as missing in the external validation. Model performance was evaluated on both the cross-validated training set and the independent testing set, using class-wise area under the receiver operating characteristic curve (AUROC), sensitivity, specificity, and F1 scores with 95% confidence intervals (95% CI). Model interpretation was performed using the Shapley Additive exPlanations (SHAP) method to assess the contribution and direction of individual taxa to model predictions^48^. SHAP provides consistent, locally accurate, and class-specific explanations by attributing prediction outputs to feature contributions based on cooperative game theory. Pareto charts and SHAP summary plots were used for visualization.

## Data Availability

The data that support the findings of this study are available for research collaboration
from the corresponding author upon reasonable request.

## Acknowledgements

This study was supported by the Clinical+ program from the Ninth People’s Hospital affiliated with the Jiao Tong University School of Medicine entitled “Discovery and validation of periodontitis biomarkers”. The funder played no role in study design, data collection, analysis and interpretation of data, or the writing of this manuscript.

## Contributions

MST, BR and YX conceived and designed the study. YX and AA analyzed the data with XY, MB, HL, and YL. YX, BR, MST drafted the manuscript with input from AR and AM. All authors have reviewed the final version of the manuscript and approved it for publication.

## Footnotes

& Online SMDI platform: https://bioinformatics.forsyth.org/smdi/index.php

