## Appendix for "Oral Rinse Sourced Microbiota in Oral Health and Disease in a Representative U.S. Adult Population"

### Supplementary Information

#### Supplementary Methods

The definition of different level of periodontitis, under CDC/AAP classification system:

(i) no periodontitis: individual absence of mild, moderate, or severe periodontitis found;

(ii) mild periodontitis: an individual with ≥2 interproximal sites with attachment loss ≥3 mm, and ≥2 interproximal sites with pocket depth ≥4 mm (not on the same tooth) or one site with pocket depth ≥5 mm;

(iii) moderate periodontitis: an individual with ≥2 interproximal sites showing attachment loss of ≥4 mm (not on the same tooth) or ≥2 interproximal sites exhibiting pocket depth of ≥5 mm (not on the same tooth);

(iv) severe periodontitis: an individual with ≥2 interproximal sites with attachment loss ≥6 mm (not on the same tooth) and ≥1 interproximal site with pocket depth ≥5 mm.

The definition of different level of periodontitis, under ACES classification system:

(i) no periodontitis: an individual with no interproximal attachment loss ≥1 mm at ≥2 non-adjacent teeth and no buccal/lingual attachment loss ≥3 mm with pocket depth >3 mm at ≥2 teeth is observed;

(ii) stage I periodontitis: an individual with interproximal attachment loss ≥1 mm at ≥2 non-adjacent teeth, with maximum attachment loss of 1-2 mm;

(iii) stage II periodontitis: an individual with interproximal attachment loss ≥1 mm at ≥2 non-adjacent teeth, with maximum attachment loss of 3-4 mm;

(iv) stage III/IV periodontitis: an individual with interproximal attachment loss ≥1 mm at ≥2 non-adjacent teeth, with maximum attachment loss ≥5 mm.

#### Supplementary Figures

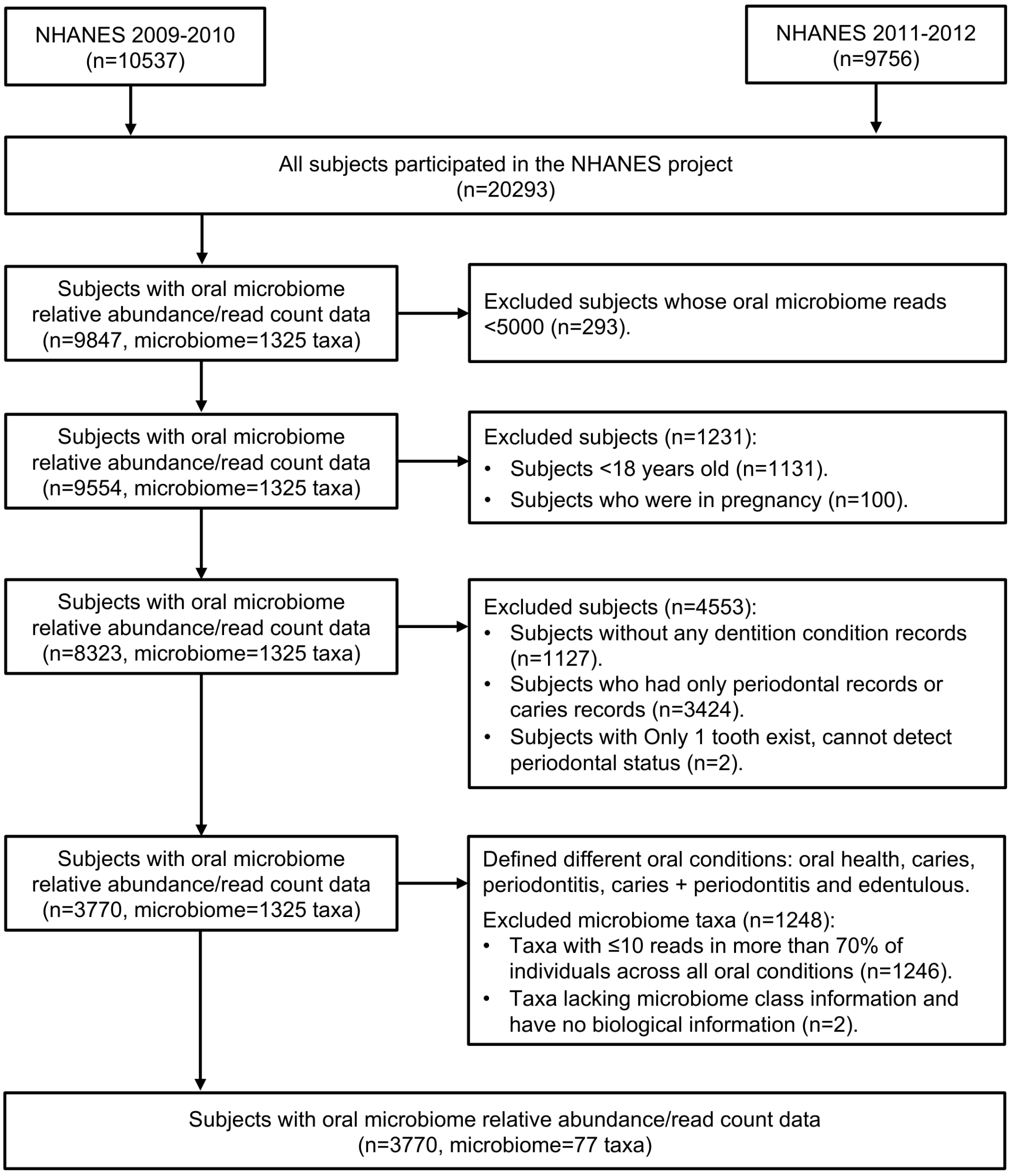


Figure S1. Flow chart of subject and microbiome inclusion process from the NHANES 2009-2010 and 2011-2012 database.

Subjects were initially identified from two NHANES cycles: 2009–2010 (n=10,537) and 2011–2012 (n=9,756), totaling 20,293 participants. Subjects with available oral microbiome data (n=9,847, with 1,325 taxa) were first selected. Sequential exclusion criteria were applied, with subjects with fewer than 5,000 microbiome reads were excluded (n = 293); subjects younger than 18 years old (n = 1,131) and those who were pregnant (n = 100) were excluded; and subjects lacking dentition records (n = 1,127), having only periodontal or only caries records (n = 3,424), or having only one remaining tooth (n = 2), which precluded periodontal assessment, were excluded.

After these steps, 3,770 participants remained with complete oral health records and high-quality oral microbiome data. Participants were classified into five oral condition categories: oral health, caries only, periodontitis only, caries + periodontitis, as well as edentulous status.

Microbial taxa were further filtered by excluding: taxa with ≤10 reads in more than 70% of individuals across all oral conditions (n = 1,246); and taxa lacking microbiome class information or known biological annotations (n = 2). The final analytical dataset included 3,770 participants and 77 microbial taxa used for further microbial analyses.

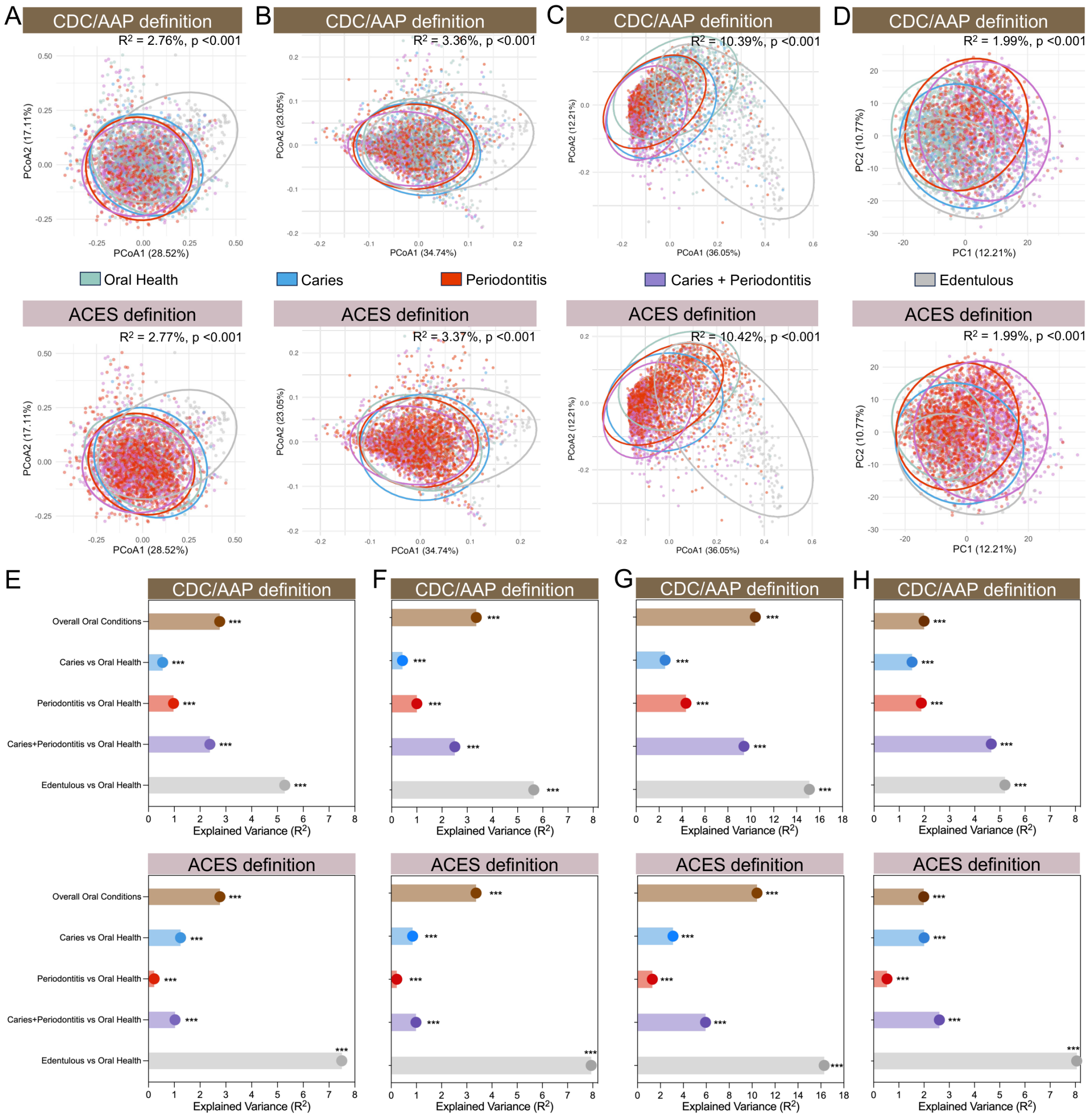


Figure S2. Variation in oral microbiota composition explained by specific oral conditions using different data analysis approaches.

(A) Principal coordinate analysis (PCoA) based on Bray-Curtis dissimilarities of all samples, colored by different oral conditions, explained variance ($R^2$) and p-values calculated by PERMANOVA test;

(B) PCoA based on weighted UniFrac dissimilarities of all samples, colored by different oral conditions, $R^2$ and p-values calculated by PERMANOVA test;

(C) PCoA based on unweighted UniFrac dissimilarities of all samples, colored by different oral conditions, $R^2$ and p-values calculated by PERMANOVA test;

(D) PCA plots based on Aitchison dissimilarities of all samples with CLR transformed data, colored by different oral conditions, $R^2$ and p-values were calculated by PERMANOVA test.

(E) Variation explained by each oral conditions defined by caries, periodontitis and dentition status, $R^2$ and p-values were calculated on relative abundance data, by PERMANOVA test based on Bray-Curtis dissimilarities;

(F) Variation explained by each oral conditions defined by caries, periodontitis and dentition status, $R^2$ and p-values were calculated on relative abundance data, by PERMANOVA test based on weighted UniFrac dissimilarities;

(G) Variation explained by each oral conditions defined by caries, periodontitis and dentition status, $R^2$ and p-values were calculated on relative abundance data, by PERMANOVA test based on unweighted UniFrac dissimilarities;

(H) Variation explained by each oral conditions defined by caries, periodontitis and dentition status, $R^2$ and p-values were calculated on CLR transformed data, by PERMANOVA test based on Aitchison dissimilarities.

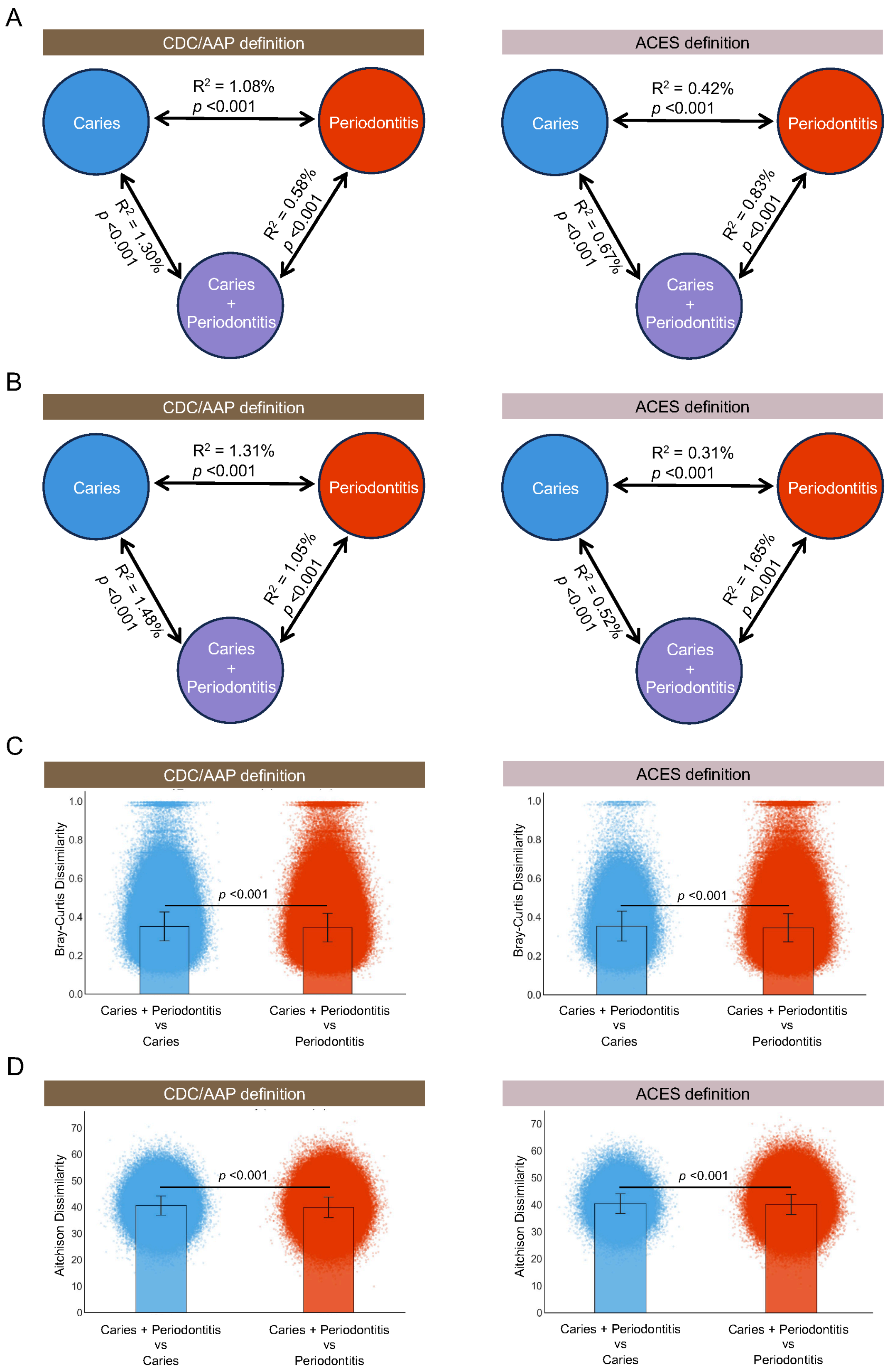


Figure S3. Inter-group variation among subjects with caries, periodontitis, and caries+periodontitis.

(A) Explained variance ($R^2$) among the three disease groups (caries, periodontitis, and caries+periodontitis), calculated from relative abundance data by PERMANOVA test based on Bray-Curtis dissimilarities; (B) E Explained variance ($R^2$) among the three disease groups, calculated from CLR-transformed data by PERMANOVA test based on Aitchison dissimilarities; (C) Pairwise comparison of dissimilarities between the comorbid group and each single disease group (caries+periodontitis vs. caries, and caries+periodontitis vs. periodontitis), based on Bray-Curtis dissimilarities of relative abundance data. Significance assessed using Mann-Whitney U test; (D) Pairwise comparison of dissimilarities between the comorbid group and each single disease group, based on Aitchison dissimilarities of CLR-transformed data. Significance assessed using Mann-Whitney U test.

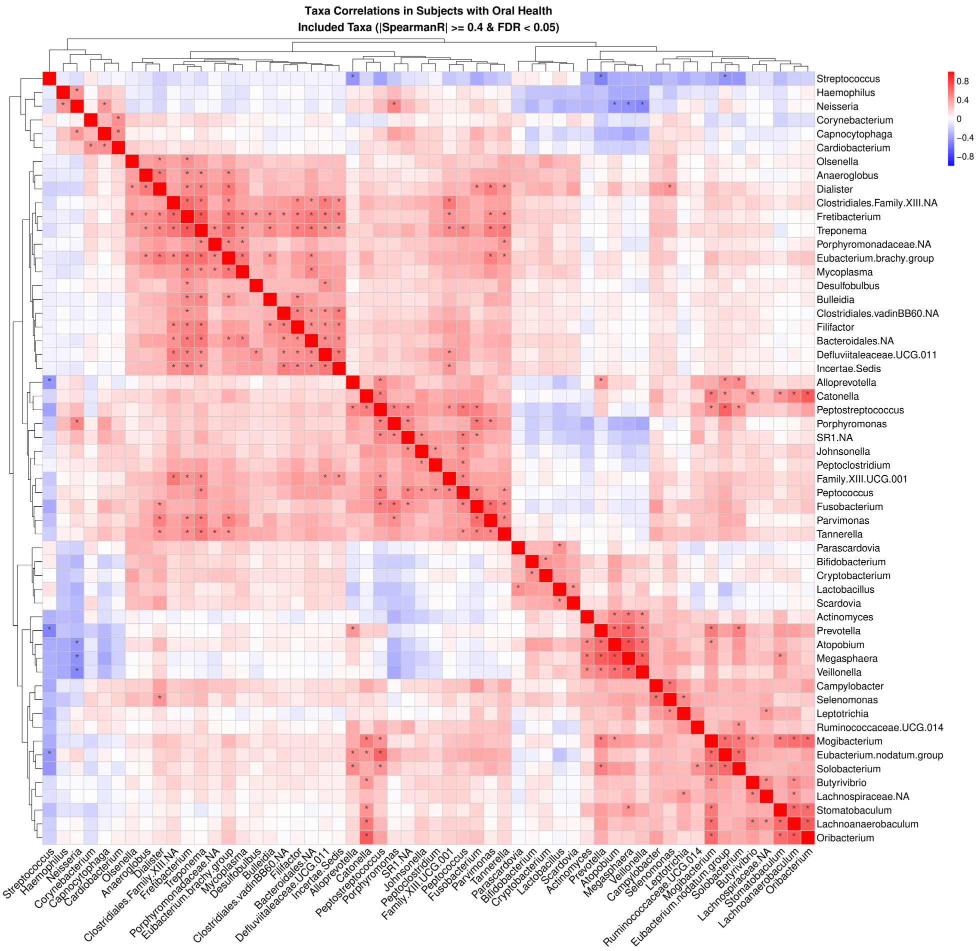


Figure S4. Taxa–taxa correlations in subjects with oral health. Correlations among microbial taxa in oral healthy individuals (without caries and periodontitis) were assessed using Spearman correlation and visualized as a hierarchical clustering heatmap. Each cell in the heatmap represents the correlation between a pair of taxa, with the color scale indicating the Spearman correlation coefficient (ρ), reflecting both the strength and direction of the correlation. Asterisks denote correlations that met the significance threshold (|ρ| ≥ 0.4 and FDR-adjusted $p < 0.05$). The heatmap is symmetrical along the diagonal, and the accompanying dendrogram illustrates the results of hierarchical clustering based on correlation patterns. The analysis accounted for the NHANES complex survey design.

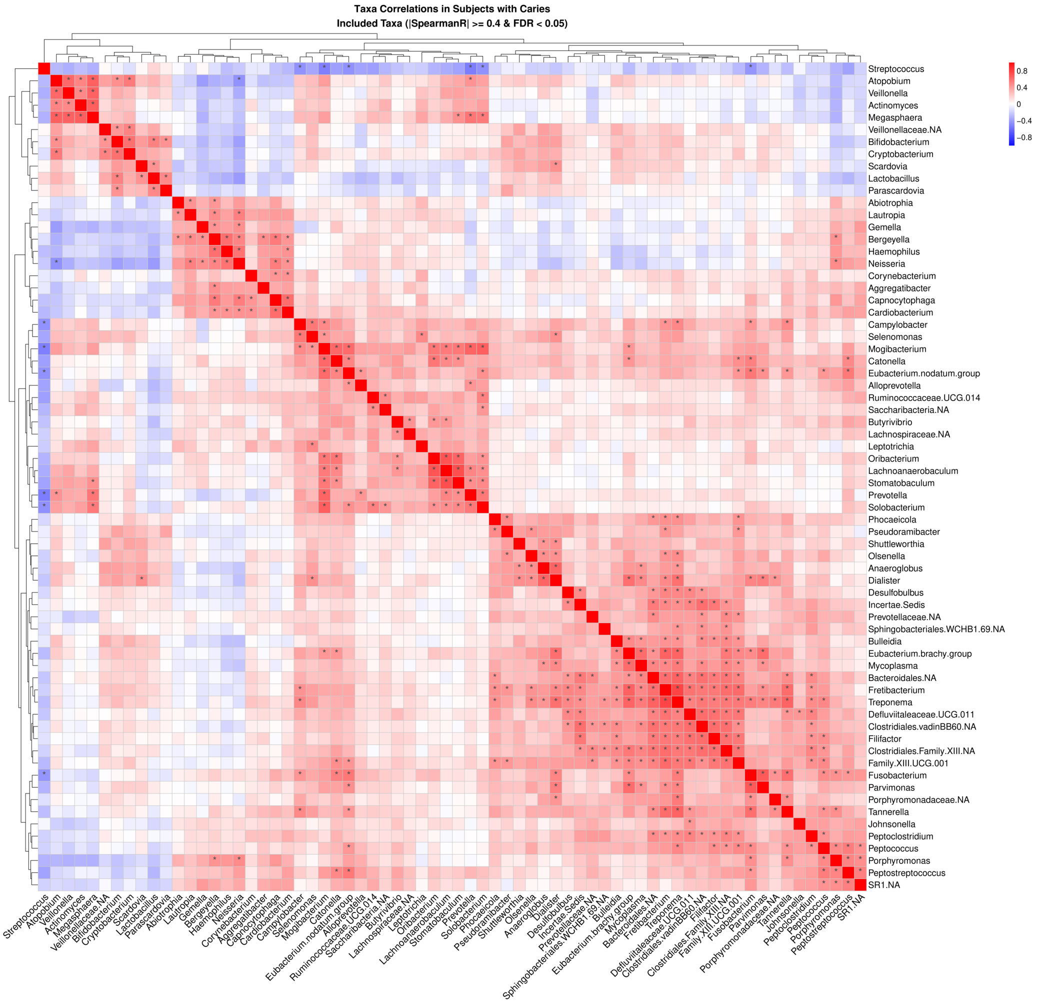


Figure S5. Taxa–taxa correlations in subjects with caries only. Correlations among microbial taxa in individuals with caries (without periodontitis) were assessed using Spearman correlation and visualized as a hierarchical clustering heatmap. Each cell in the heatmap represents the correlation between a pair of taxa, with the color scale indicating the Spearman correlation coefficient (ρ), reflecting both the strength and direction of the correlation. Asterisks denote correlations that met the significance threshold (|ρ| value ≥ 0.4 and FDR-adjusted $p < 0.05$). The heatmap is symmetrical along the diagonal, and the accompanying dendrogram illustrates the results of hierarchical clustering based on correlation patterns. The analysis accounted for the NHANES complex survey design.

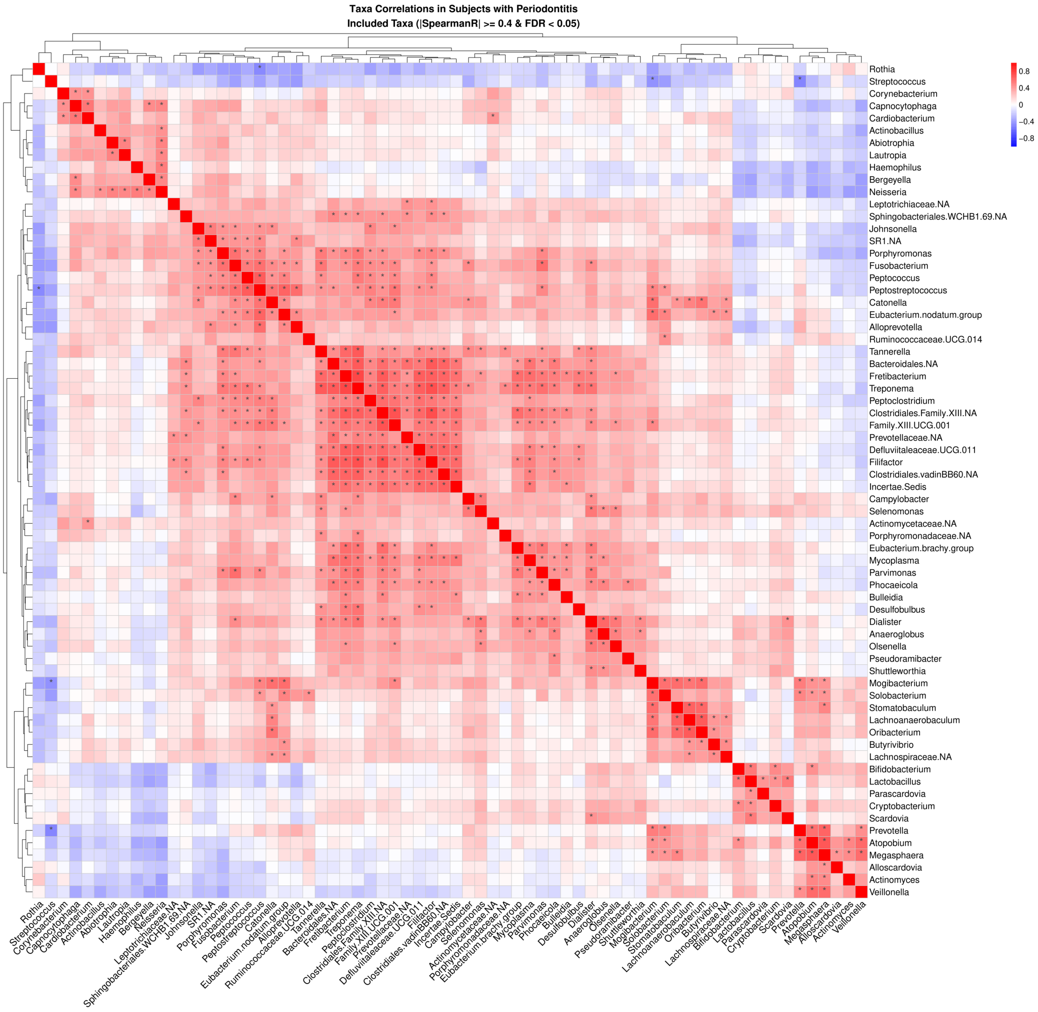


Figure S6. Taxa–taxa correlations in subjects with periodontitis only. Correlations among microbial taxa in individuals with periodontitis (without caries) were assessed using Spearman correlation and visualized as a hierarchical clustering heatmap. Each cell in the heatmap represents the correlation between a pair of taxa, with the color scale indicating the Spearman correlation coefficient (ρ), reflecting both the strength and direction of the correlation. Asterisks denote correlations that met the significance threshold (|ρ| value ≥ 0.4 and FDR-adjusted $p < 0.05$). The heatmap is symmetrical along the diagonal, and the accompanying dendrogram illustrates the results of hierarchical clustering based on correlation patterns. The analysis accounted for the NHANES complex survey design.

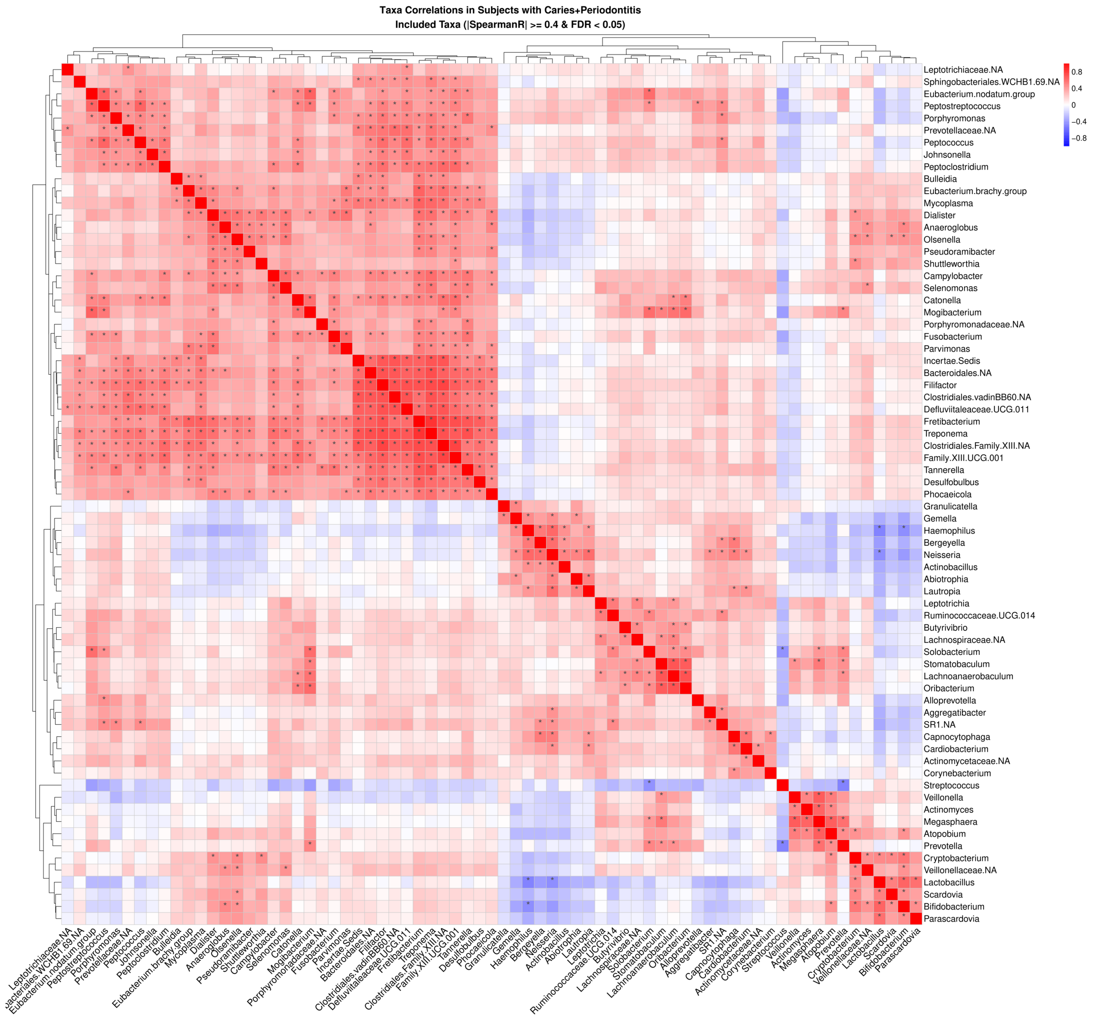


Figure S7. Taxa–taxa correlations in subjects with caries+periodontitis. Correlations among microbial taxa in individuals with caries+periodontitis were assessed using Spearman correlation and visualized as a hierarchical clustering heatmap. Each cell in the heatmap represents the correlation between a pair of taxa, with the color scale indicating the Spearman correlation coefficient (ρ), reflecting both the strength and direction of the correlation. Asterisks denote correlations that met the significance threshold (|ρ| value ≥ 0.4 and FDR-adjusted $p < 0.05$). The heatmap is symmetrical along the diagonal, and the accompanying dendrogram illustrates the results of hierarchical clustering based on correlation patterns. The analysis accounted for the NHANES complex survey design.

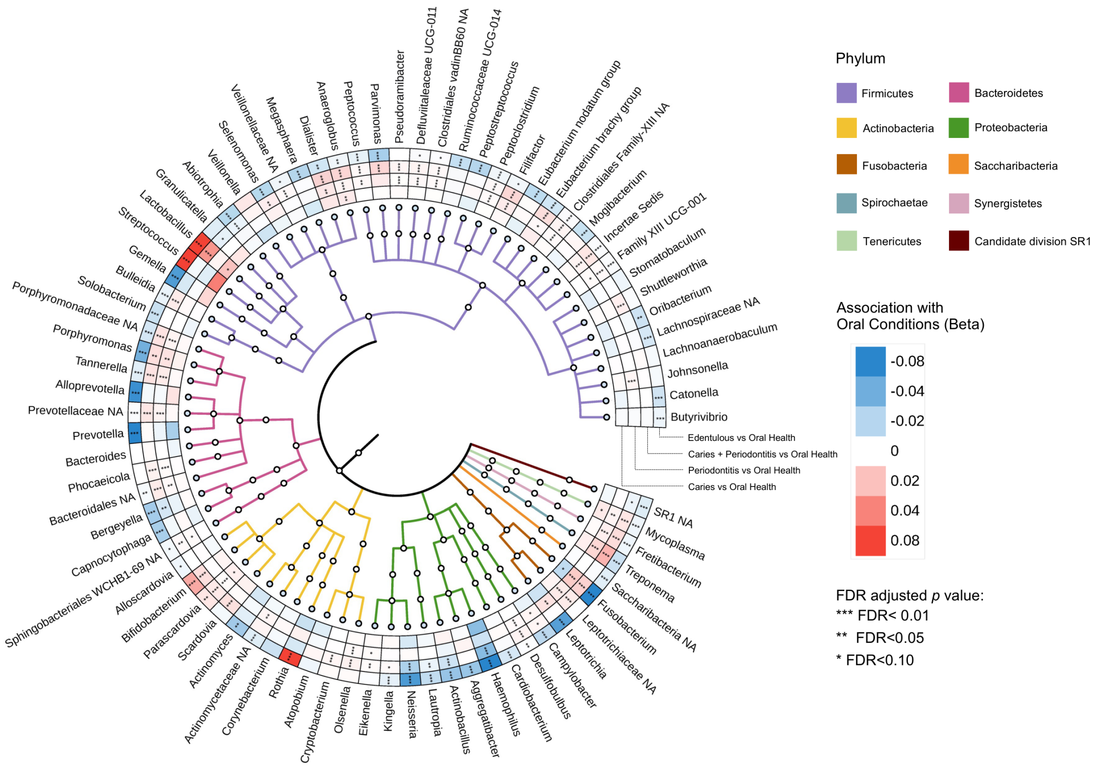


Figure S8. Identification of oral condition-associated microbial taxa.

This analysis assessed significant associations between oral microbial taxa and different oral conditions under the ACES periodontitis definitions in the NHANES dataset. Relative abundance data were normalized using arcsine square root (Arrcsin-Sqrt) transformation prior to analysis. Cross-sectional associations were evaluated using linear regression model, adjusting for sex, body mass index (BMI), income-to-poverty ratio, education level and systematic diseases of diabetes and hypertension. The analysis accounted for the NHANES complex survey design. Multiple comparisons were corrected using the false discovery rate (FDR) method with a target rate of 0.10. In the heatmap, color intensity represents the strength and direction of associations between taxa and oral conditions (each compared to oral health). Colors on the clade of the phylogenetic tree denote different bacterial phyla.

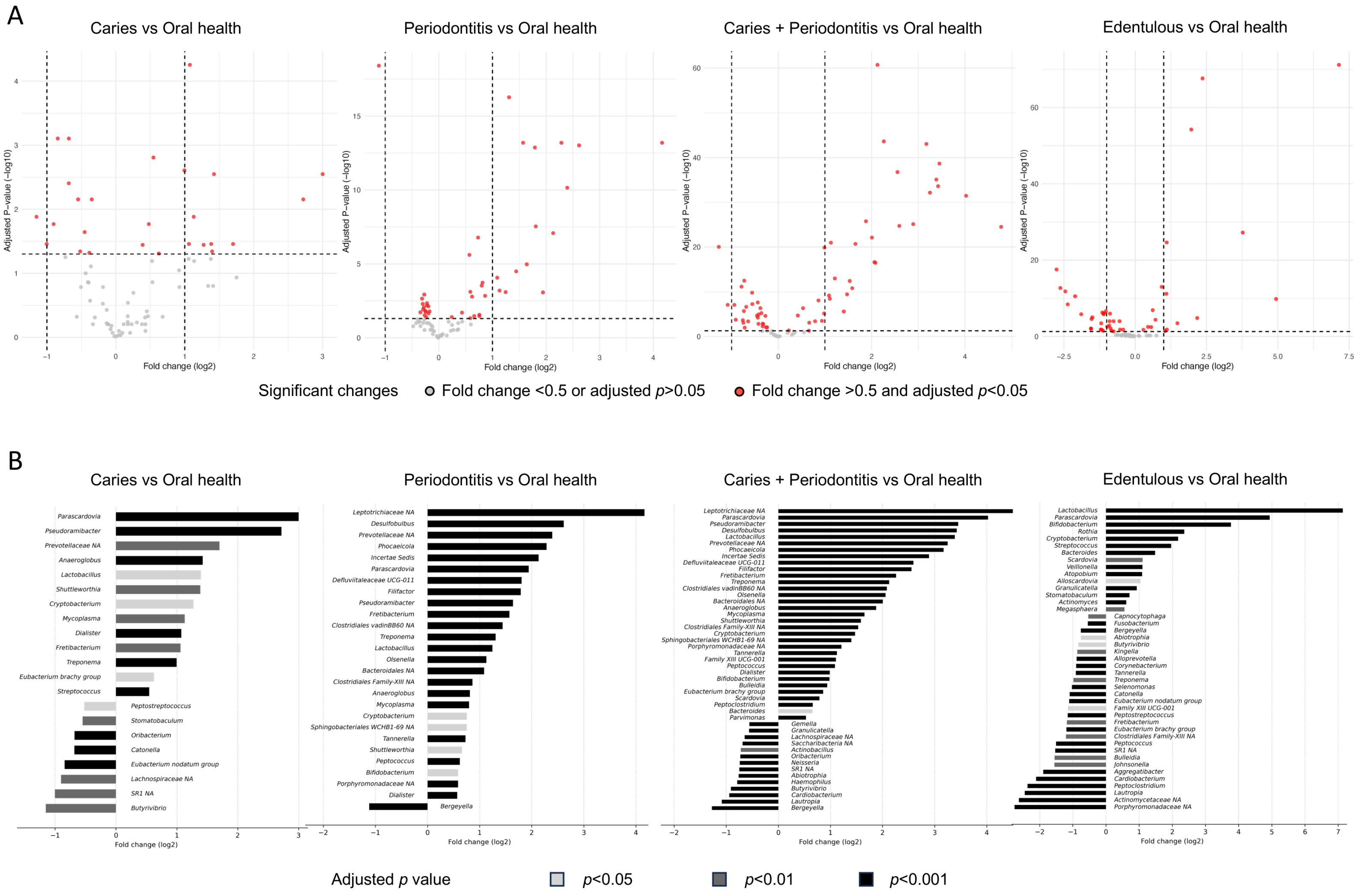
A
Caries vs Oral health
Periodontitis vs Oral health
Caries + Periodontitis vs Oral health
Edentulous vs Oral health
Adjusted P−value (−log10)
Fold change (log2)
Significant changes
Fold change <0.5 or adjusted p>0.05
Fold change >0.5 and adjusted p<0.05
B
Caries vs Oral health
Periodontitis vs Oral health
Caries + Periodontitis vs Oral health
Edentulous vs Oral health
Fold change (log2)
Adjusted p value
p<0.05
p<0.01
p<0.001

Figure S9. Differential abundance of taxa across oral conditions analyzed through DESeq2 based on the ACES periodontitis definition.

(A) Volcano plots displaying differential abundance comparisons between each oral condition and oral health. Taxa with an adjusted *p* value < 0.05 and absolute $\log_2$ fold change > 0.5 are highlighted in red.

(B) Bar plots presenting significantly altered taxa across oral conditions, with bar colors indicating different levels of statistical significance based on adjusted *p* values.

Comparisons were performed using DESeq2 on raw read count data, followed by false discovery rate (FDR) correction for multiple testing. $\text{Log}_2$ fold changes estimated by the DESeq2 model were directly used for visualization in volcano and bar plots. Oral conditions compared to oral health included caries only, periodontitis only, combined caries and periodontitis, and edentulous status.

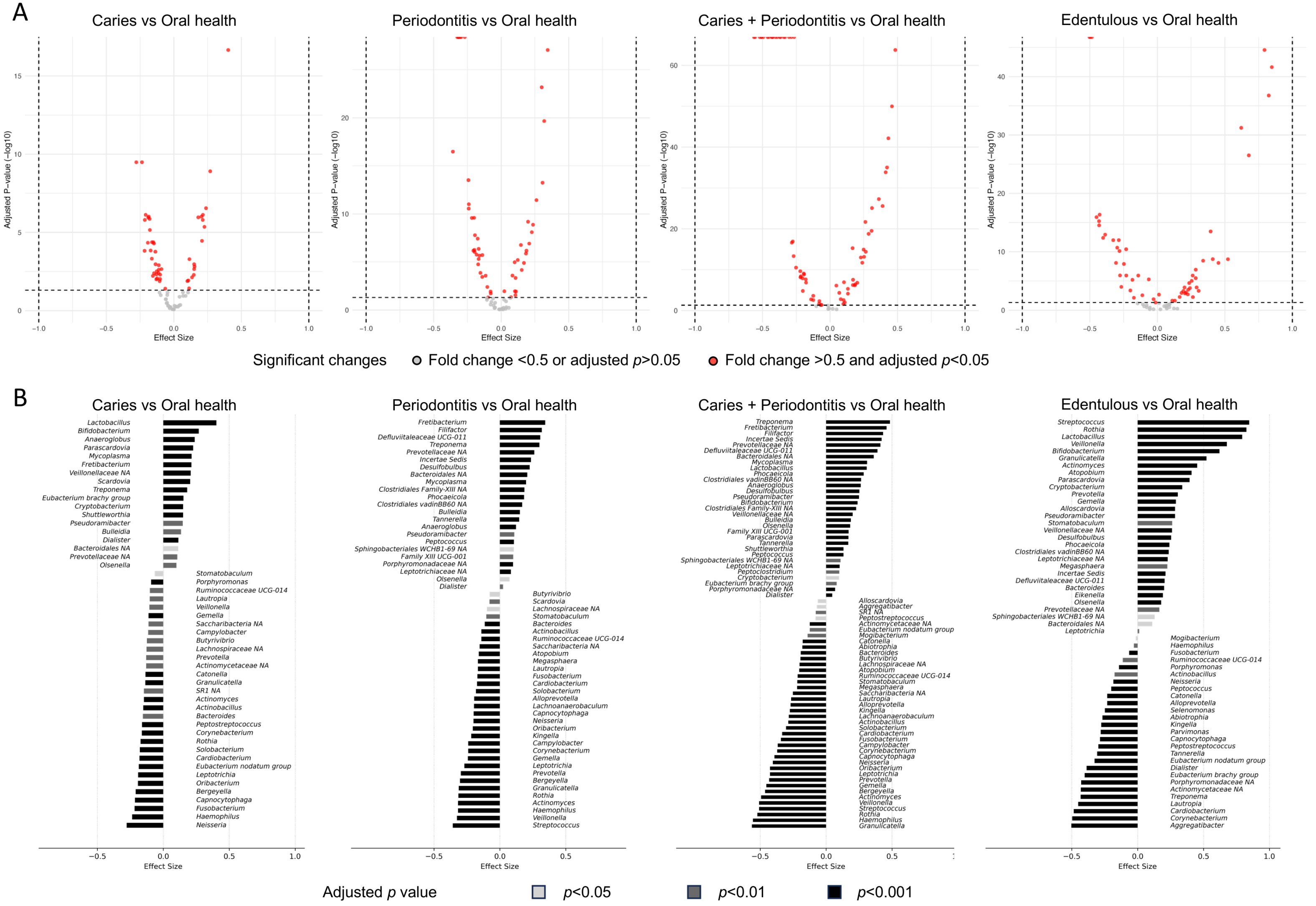
A
Caries vs Oral health
Periodontitis vs Oral health
Caries + Periodontitis vs Oral health
Edentulous vs Oral health
Adjusted P−value (−log10)
Effect Size
Significant changes
Fold change <0.5 or adjusted p>0.05
Fold change >0.5 and adjusted p<0.05
B
Caries vs Oral health
Periodontitis vs Oral health
Caries + Periodontitis vs Oral health
Edentulous vs Oral health
Effect Size
Adjusted p value
p<0.05
p<0.01
p<0.001

Figure S10. Differential abundance of taxa across oral conditions analyzed through ALDEx2 based on the ACES periodontitis definition.

(A) Volcano plots displaying differential abundance comparisons between each oral condition and oral health. Taxa with an adjusted *p* value < 0.05 are highlighted in red.

(B) Bar plots presenting significantly altered taxa across oral conditions, with bar colors indicating different levels of statistical significance based on adjusted *p* values.

Comparisons were performed using ALDEx2, which applies a centered log-ratio (CLR) transformation of the count data with Monte Carlo sampling from the Dirichlet distribution, followed by false discovery rate (FDR) correction for multiple testing. The estimated effect size, representing the between-group difference in CLR abundance, was directly used for visualization in volcano and bar plots. Oral conditions compared to oral health included caries only, periodontitis only, combined caries and periodontitis, and edentulous status.

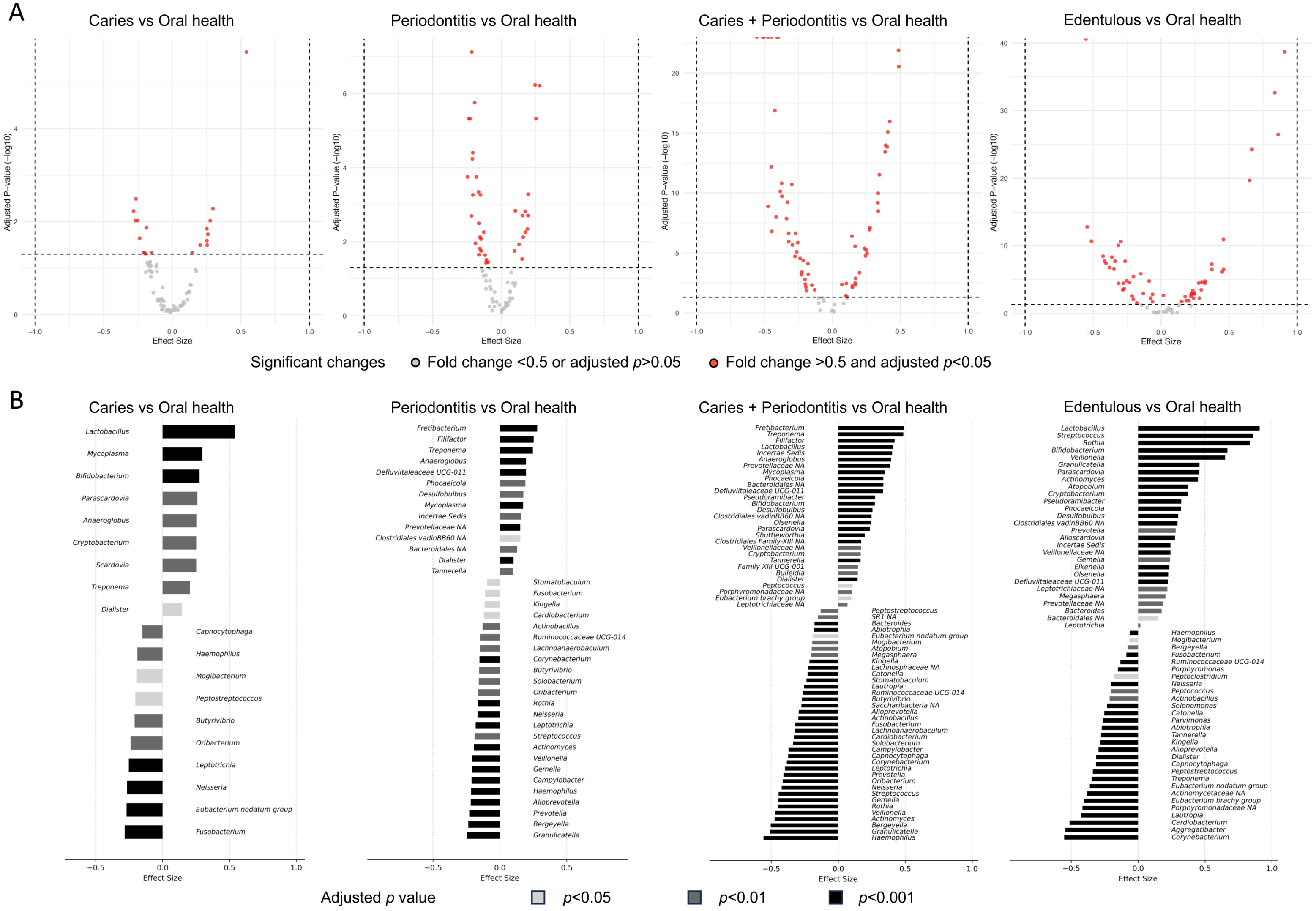
A
Caries vs Oral health
Periodontitis vs Oral health
Caries + Periodontitis vs Oral health
Edentulous vs Oral health
Adjusted P-value (−log10)
Effect Size
Significant changes
Fold change <0.5 or adjusted p>0.05
Fold change >0.5 and adjusted p<0.05
B
Caries vs Oral health
Lactobacillus
Mycoplasma
Bifidobacterium
Parascardovia
Anaeroglobus
Cryptobacterium
Scardovia
Treponema
Dialister
Capnocytophaga
Haemophilus
Mogibacterium
Peptostreptococcus
Butyrivibrio
Oribacterium
Leptotrichia
Neisseria
Eubacterium nodatum group
Fusobacterium
Periodontitis vs Oral health
Fretibacterium
Filifactor
Treponema
Anaeroglobus
Defluviitaleaceae UCG-011
Phocaeicola
Desulfobulbus
Mycoplasma
Incertae Sedis
Prevotellaceae NA
Clostridiales vadinBB60 NA
Bacteroidales NA
Dialister
Tannerella
Stomatobaculum
Fusobacterium
Kingella
Cardiobacterium
Actinobacillus
Ruminococcaceae UCG-014
Lachnoanaerobaculum
Corynebacterium
Butyrivibrio
Solobacterium
Oribacterium
Rothia
Neisseria
Leptotrichia
Streptococcus
Actinomyces
Veillonella
Gemella
Campylobacter
Haemophilus
Alloprevotella
Prevotella
Bergeyella
Granulicatella
Caries + Periodontitis vs Oral health
Fretibacterium
Treponema
Filifactor
Lactobacillus
Incertae Sedis
Anaeroglobus
Prevotellaceae NA
Mycoplasma
Phocaeicola
Bacteroidales NA
Defluviitaleaceae UCG-011
Pseudoramibacter
Bifidobacterium
Desulfobulbus
Clostridiales vadinBB60 NA
Olsenella
Parascardovia
Shuttleworthia
Clostridiales Family-XIII NA
Veillonellaceae NA
Cryptobacterium
Tannerella
Family XIII UCG-001
Bulleidia
Dialister
Peptococcus
Porphyromonadaceae NA
Eubacterium brachy group
Leptotrichiaceae NA
Peptostreptococcus
SR1 NA
Bacteroides
Abiotrophia
Eubacterium nodatum group
Mogibacterium
Atopobium
Megasphaera
Kingella
Lachnospiraceae NA
Catonella
Stomatobaculum
Lautropia
Ruminococcaceae UCG-014
Butyrivibrio
Saccharibacteria NA
Alloprevotella
Actinobacillus
Fusobacterium
Lachnoanaerobaculum
Cardiobacterium
Solobacterium
Campylobacter
Capnocytophaga
Corynebacterium
Leptotrichia
Prevotella
Oribacterium
Neisseria
Streptococcus
Gemella
Rothia
Veillonella
Actinomyces
Bergeyella
Granulicatella
Haemophilus
Edentulous vs Oral health
Lactobacillus
Streptococcus
Rothia
Bifidobacterium
Veillonella
Granulicatella
Parascardovia
Actinomyces
Atopobium
Cryptobacterium
Pseudoramibacter
Phocaeicola
Desulfobulbus
Clostridiales vadinBB60 NA
Prevotella
Alloscardovia
Incertae Sedis
Veillonellaceae NA
Gemella
Eikenella
Olsenella
Defluviitaleaceae UCG-011
Leptotrichiaceae NA
Megasphaera
Prevotellaceae NA
Bacteroides
Bacteroidales NA
Leptotrichia
Haemophilus
Mogibacterium
Bergeyella
Fusobacterium
Ruminococcaceae UCG-014
Porphyromonas
Peptoclostridium
Neisseria
Peptococcus
Actinobacillus
Selenomonas
Catonella
Parvimonas
Abiotrophia
Tannerella
Kingella
Alloprevotella
Dialister
Capnocytophaga
Peptostreptococcus
Treponema
Eubacterium nodatum group
Actinomycetaceae NA
Eubacterium brachy group
Porphyromonadaceae NA
Lautropia
Cardiobacterium
Aggregatibacter
Corynebacterium
Effect Size
Adjusted p value
p<0.05
p<0.01
p<0.001

Figure S11. Differential abundance of taxa across oral conditions analyzed through ALDEx2 based on the ACES periodontitis definition.

(A) Volcano plots displaying differential abundance comparisons between each oral condition and oral health. Taxa with an adjusted *p* value < 0.05 are highlighted in red.

(B) Bar plots presenting significantly altered taxa across oral conditions, with bar colors indicating different levels of statistical significance based on adjusted *p* values.

Comparisons were performed using ALDEx2, which applies a centered log-ratio (CLR) transformation of the count data with Monte Carlo sampling from the Dirichlet distribution, followed by false discovery rate (FDR) correction for multiple testing. The estimated effect size, representing the between-group difference in CLR abundance, was directly used for visualization in volcano and bar plots. Oral conditions compared to oral health included caries only, periodontitis only, combined caries and periodontitis, and edentulous status.

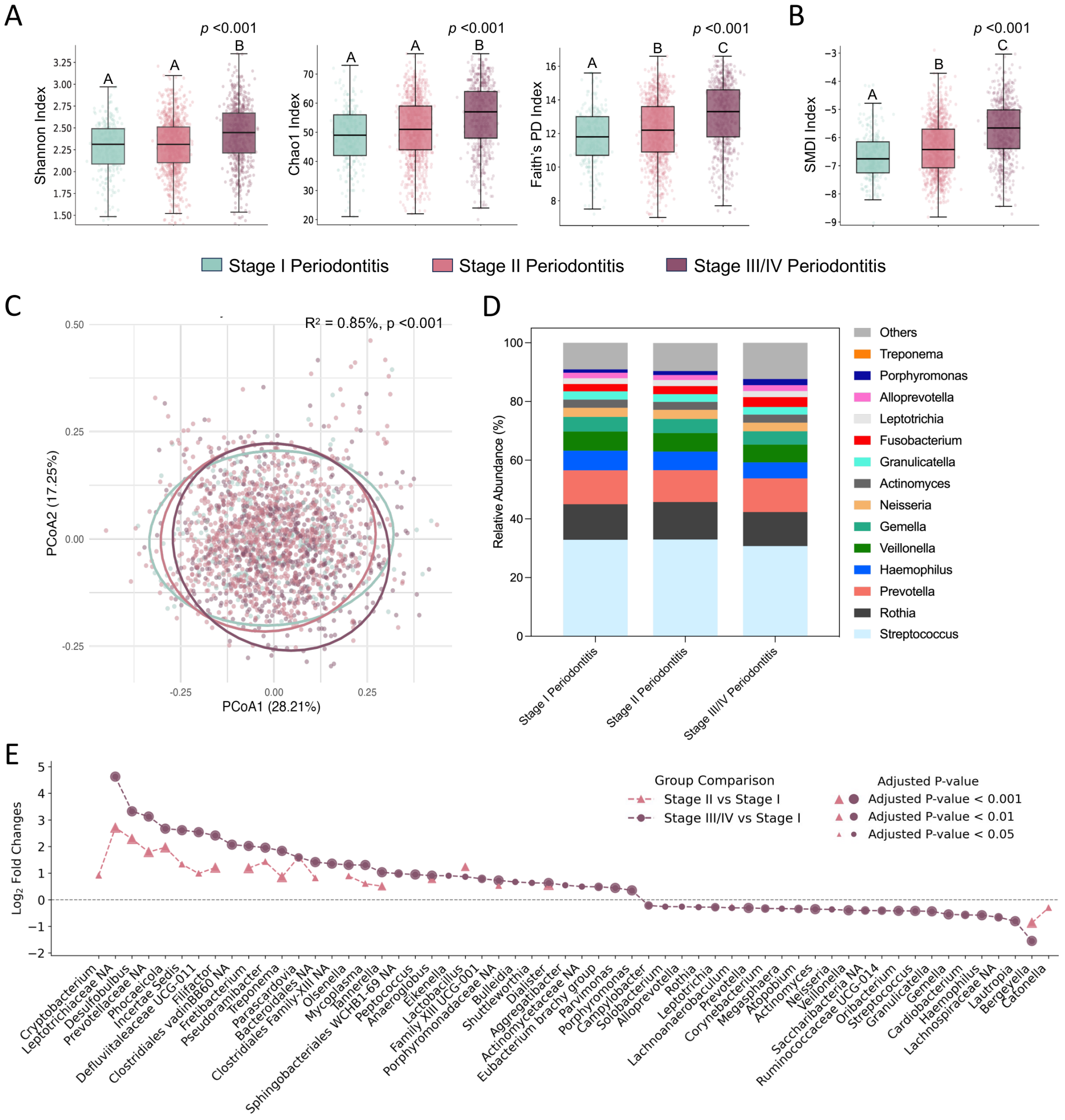


Figure S12. Dose-dependent association between oral microbiome profiles and periodontitis severity under the ACES definition.

(A) Alpha diversity comparisons across periodontitis severity status, measured by Shannon, Chao1, and Faith's PD indices;

(B) Changes in periodontitis-related microbiome dysbiosis across periodontitis severity status, measured by the subgingival microbial dysbiosis index (SMDI);

(C) Principal coordinate analysis (PCoA) plot based on Bray-Curtis dissimilarities of relative abundance data, comparing microbial community structures across periodontitis severity status. Explained variance ($R^2$) and *p* values were calculated by PERMANOVA test;

(D) Average relative abundances of the most dominant taxa (>1% average relative abundance) across periodontitis severity status;

(E) Differential abundance comparisons between relatively healthy periodontal status (stage I periodontitis in ACES definition) and more advanced periodontitis stages. Each dot represents a taxon with significant changes, with dot position indicating the magnitude of change ($log_2$-transformed fold change) and dot size reflecting adjusted *p* value significance. Comparisons of the differential abundance were performed using the DESeq2 analysis on read count data, followed by false discovery rate (FDR) correction for multiple testing. Taxa with adjusted $p$ value < 0.05 underwent $log_2$ transformation for visualization.

For (A) and (B), *p* values represent Kruskal-Wallis test results across all groups. Post-hoc pairwise comparisons were conducted using Dunn’s test, with different letters above bars indicate statistically significant differences ($p < 0.05$), while identical letters indicate no significant difference. All statistical analyses accounted for the NHANES complex survey design.

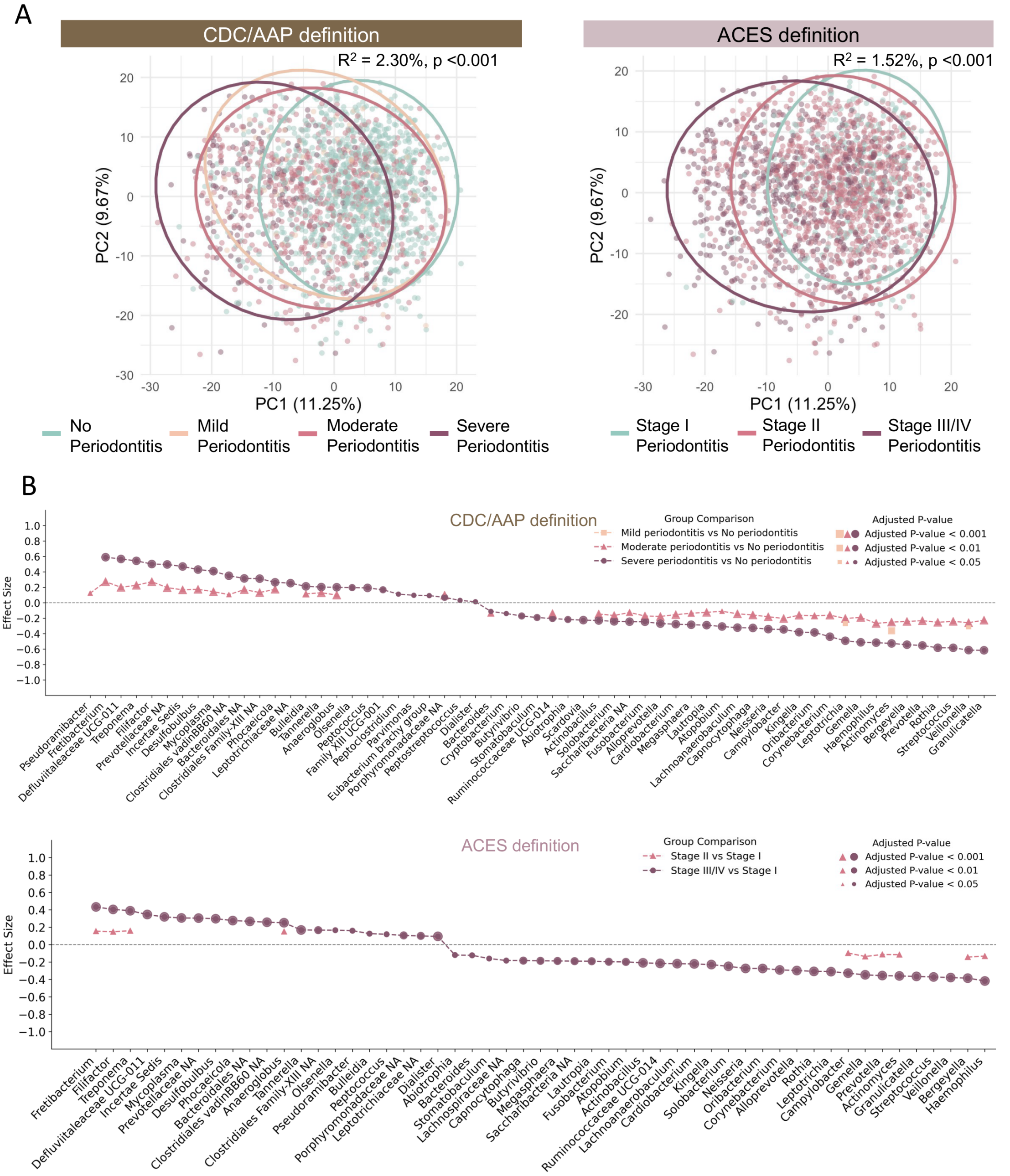


Figure S13. Dose-dependent associations between oral microbiome profiles and periodontitis severity analyzed using CLR-transformed data.

(A) Principal component analysis (PCA) plots based on Aitchison dissimilarities of CLR-transformed data, colored by periodontitis severity status. Explained variance ($R^2$) and *p* values were calculated by PERMANOVA.

(B) Differential abundance comparisons between relatively healthy periodontal status (no periodontitis under CDC/AAP or Stage I periodontitis under ACES) and more advanced stages of periodontitis. Each dot represents a taxon with significant differences, the dot position reflects the magnitude of change (effect size estimated from between-group CLR differences), and the dot size indicates the level of statistical significance (adjusted p-value). Statistical significance was assessed using Welch's t-test and Wilcoxon rank test on Monte Carlo samples generated by ALDEx2, followed by false discovery rate (FDR) correction. Only taxa with adjusted p-values < 0.05 are shown in the plot.

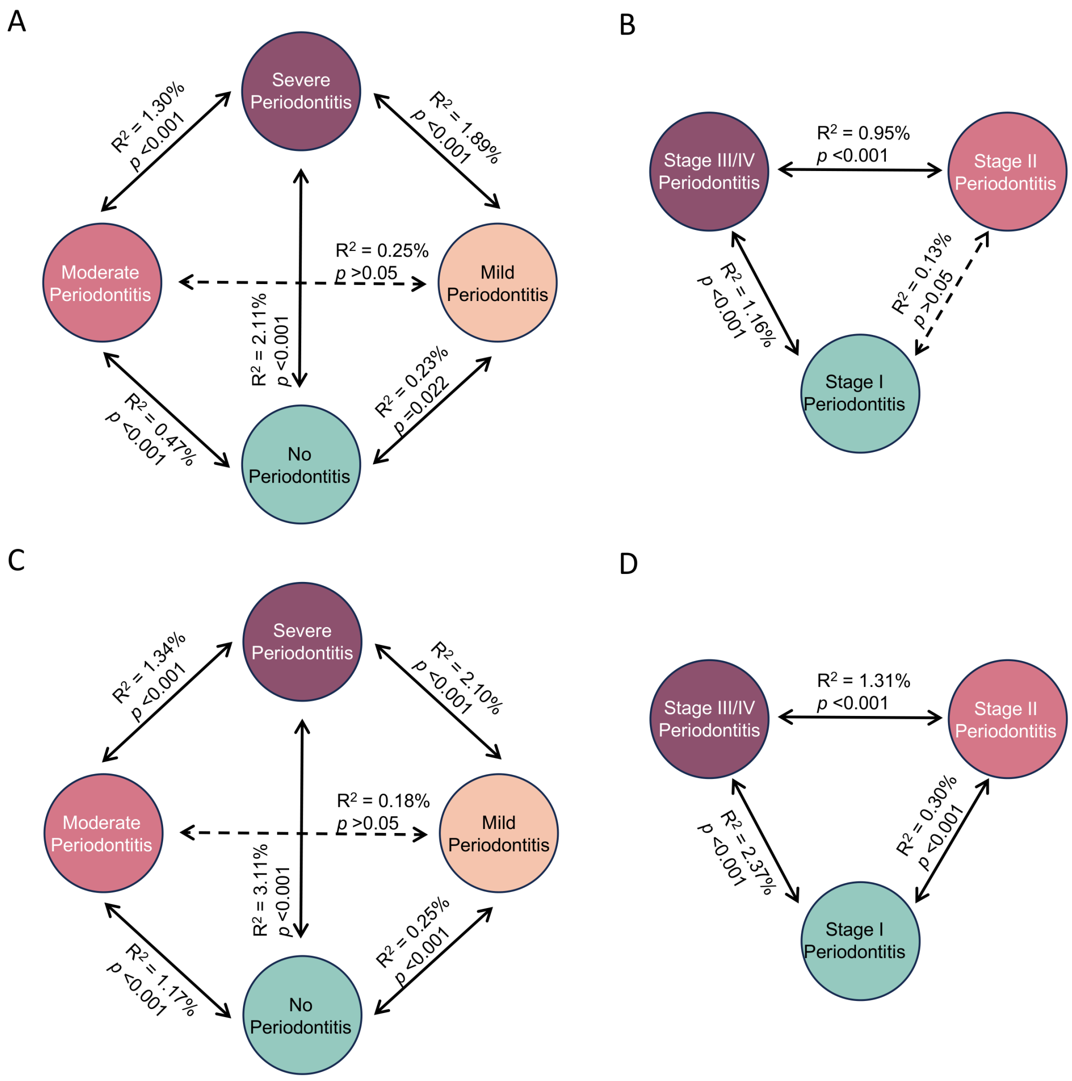


Figure S14. Inter-group variation among subjects with different severities of periodontitis, based on CDC/AAP definition and ACES definition.

(A) Explained variance ($R^2$) among the four periodontitis severity groups as defined by CDC/AAP definition (no periodontitis, mild periodontitis, moderate periodontitis, severe periodontitis), calculated from relative abundance data by PERMANOVA test based on Bray-Curtis dissimilarities; (B) Explained variance ($R^2$) among the three periodontitis severity groups as defined by ACES definition (stage I periodontitis, stage II periodontitis, stage III/IV periodontitis), calculated from relative abundance data by PERMANOVA test based on Bray-Curtis dissimilarities; (C) Explained variance ($R^2$) among the four periodontitis severity groups as defined by CDC/AAP definition, calculated from CLR-transformed data by PERMANOVA test based on Aitchison dissimilarities; (D) Explained variance ($R^2$) among the three periodontitis severity groups as defined by ACES definition, calculated from CLR-transformed data by PERMANOVA test based on Aitchison dissimilarities. Dotted lines indicate pairwise comparisons with no statistically significant differences, while solid lines indicate statistically significant differences between groups.

A

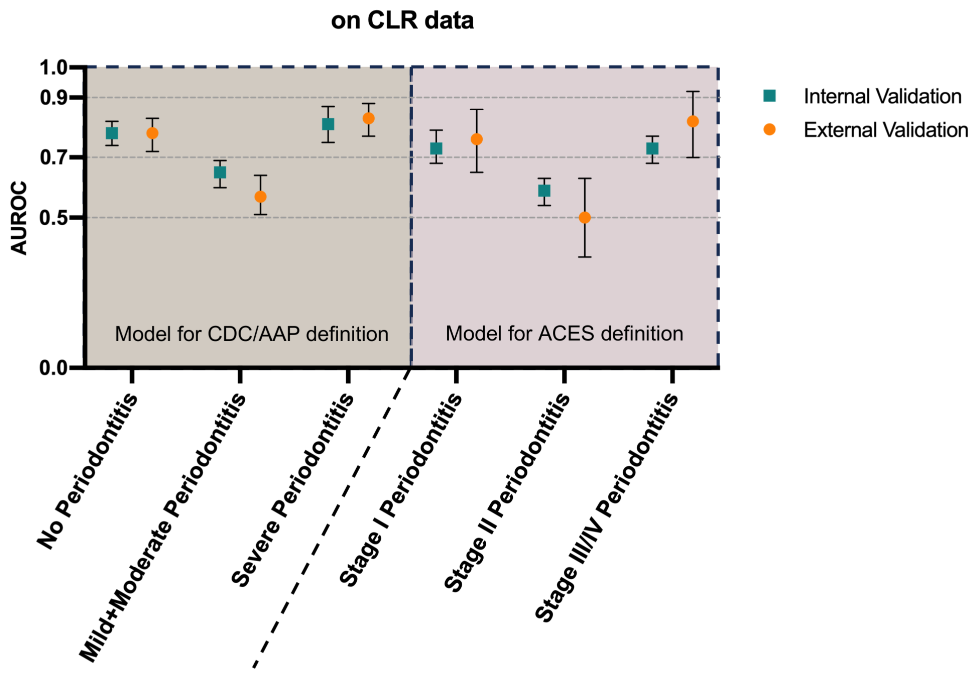


B

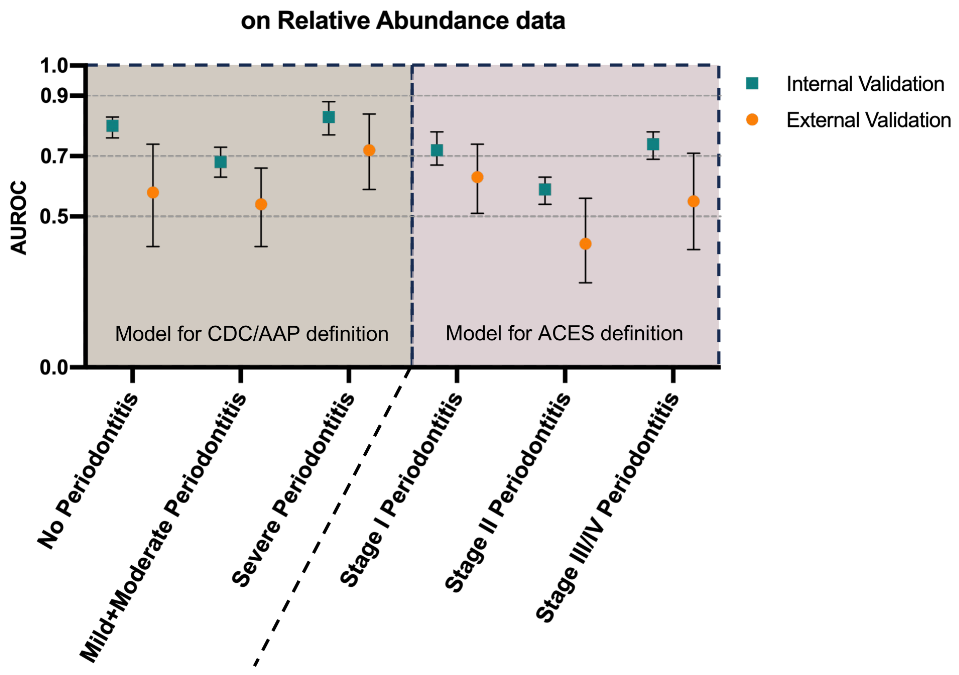


Figure S15. Comparison of machine learning model performance between internal and external validation.

(A) Comparison of AUROC between internal and external validation for the optimal random forest model developed on CLR-transformed data from the NHANES database, using either the CDC/AAP or ACES definition of periodontitis. Internal validation was conducted with a leave-one-dataset-out approach within NHANES, and external validation was performed using a local cohort established by the researchers.

(B) Comparison of AUROC between internal and external validation for the optimal random forest model developed on relative abundance data from the NHANES database, using either the CDC/AAP or ACES definition of periodontitis. Internal validation was conducted with a leave-one-dataset-out approach within NHANES, and external validation was performed using a local cohort established by the researchers.

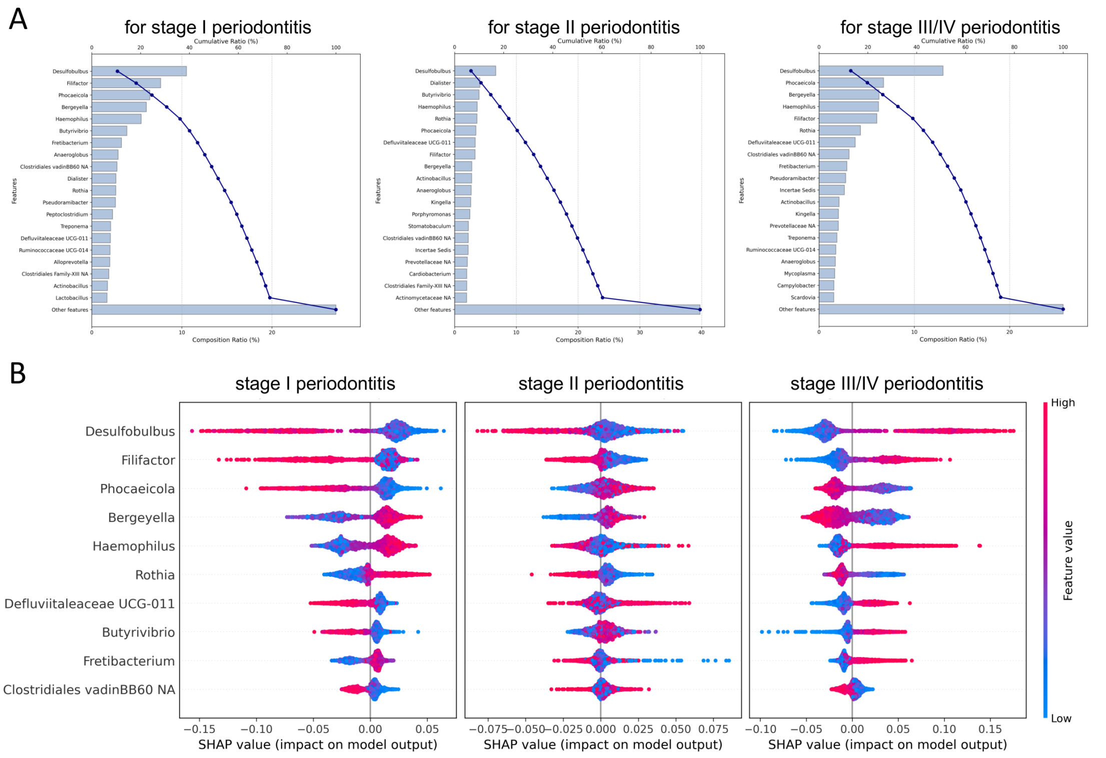


Figure S16. Periodontitis-related taxa interpreted by machine learning on CLR-transformed relative abundance data across different periodontitis severity stutas defined by the ACES definition.

(A) Pareto plots showing the single and cumulative contribution of dominant taxa to model interpretability, as determined by the random forest multi-class model;

(B) Shapley additive explanation (SHAP) summary plots for the top10 taxa contributing to model interpretability. Each point represents a sample, colored by the CLR transformed relative abundance of the corresponding taxon (blue to red representing low to high abundance). The x-axis shows the SHAP value, indicating both the magnitude and direction of each taxon's impact on the model output.

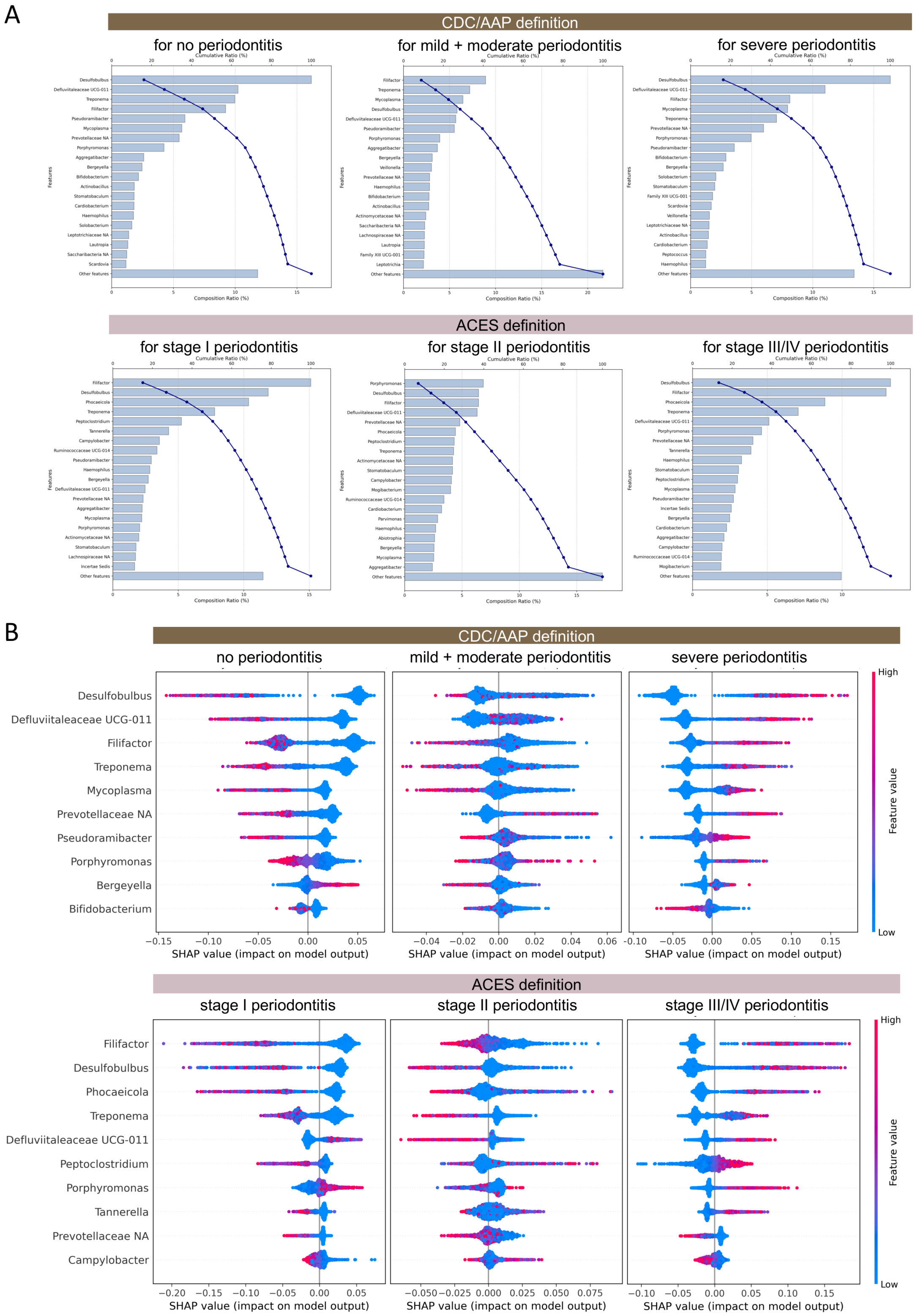


Figure S17. Periodontitis-associated taxa identified by machine learning on relative abundance data across different periodontitis severity status defined by the CDC/AAP and ACES definition.

(A) Pareto plots showing the single and cumulative contribution of dominant taxa to model interpretability, as determined by the random forest multi-class model;

(B) Shapley additive explanation (SHAP) summary plots for the top10 taxa contributing to model interpretability. Each point represents a sample, colored by the relative abundance of the corresponding taxon (blue to red representing low to high relative abundance). The x-axis shows the SHAP value, indicating both the magnitude and direction of each taxon's impact on the model output.

**Supplementary Tables**

Table S1. Characteristics of study participants with different oral health conditions based on caries and periodontitis status with CDC/AAP periodontitis definitions.

| **CDC/AAP definition** | **Overall** | **Edentulous** | **Oral Health** | **Caries** | **Periodontitis** | **Caries+Periodontitis** | ***p* value** |
|---|---|---|---|---|---|---|---|
| **N** | 3770 | 293 | 1056 | 440 | 877 | 1104 | |
| Age | 49 (39-59) | 61 (54-65) | 44 (36-54) | 42 (35-50) | 53 (44-62) | 51 (41-59) | <0.001 |
| Sex | | | | | | | |
| Male | 1955 (51.9%) | 154 (52.6%) | 417 (39.5%) | 179 (40.7%) | 501 (57.1%) | 704 (63.8%) | <0.001 |
| Female | 1815 (48.1%) | 139 (47.4%) | 639 (60.5%) | 261 (59.3%) | 376 (42.9%) | 400 (36.2%) | |
| BMI (kg/$m^2$) | 28.3 (24.7-32.9) | 27.7 (24.5-33.1) | 27.3 (24.0-31.8) | 29.3 (25.4-33.8) | 28.4 (25.0-32.9) | 28.7 (25.0-33.4) | <0.001 |
| Income-poverty ratio | 1.8 (1.0-4.0) | 1.1 (0.8-2.0) | 3.6 (1.6-5.0) | 1.6 (0.8-3.5) | 2.0 (1.1-4.1) | 1.2 (0.8-2.3) | <0.001 |
| Education level | | | | | | | |
| Less than 9th grade | 433 (11.5%) | 56 (19.1%) | 46 (4.4%) | 32 (7.3%) | 98 (11.2%) | 201 (18.2%) | <0.001 |
| 9-11th grade | 632 (16.8%) | 84 (28.7%) | 86 (8.1%) | 71 (16.1%) | 117 (13.3%) | 274 (24.8%) | |

| | | | | | | | |
|---|---|---|---|---|---|---|---|
| High school grad/GED or Equivalent | 834 (22.1%) | 73 (24.9%) | 140 (13.3%) | 126 (28.6%) | 213 (24.3%) | 282 (25.5%) | |
| Some college or AA degree | 1013 (26.9%) | 64 (21.8%) | 294 (27.8%) | 147 (33.4%) | 253 (28.8%) | 255 (23.1%) | |
| College graduate or above | 856 (22.7%) | 16 (5.5%) | 490 (46.4%) | 63 (14.3%) | 195 (22.2%) | 92 (8.3%) | |
| Missing | 2 (0.1%) | 0 (0.0%) | 0 (0.0%) | 1 (0.2%) | 1 (0.1%) | 0 (0.0%) | |
| Alcohol consumption | | | | | | | |
| Never | 687 (18.2%) | 103 (35.2%) | 112 (10.6%) | 81 (18.4%) | 155 (17.7%) | 236 (21.4%) | |
| Everyday/drink | 158 (4.2%) | 11 (3.8%) | 25 (2.4%) | 15 (3.4%) | 44 (5%) | 63 (5.7%) | |
| Every week/drink | 930 (24.7%) | 41 (14%) | 309 (29.3%) | 98 (22.3%) | 214 (24.4%) | 268 (24.3%) | <0.001 |
| Every month/drink | 532 (14.1%) | 28 (9.6%) | 192 (18.2%) | 57 (13%) | 117 (13.3%) | 138 (12.5%) | |
| Several month/drink | 677 (18%) | 57 (19.5%) | 193 (18.3%) | 96 (21.8%) | 162 (18.5%) | 169 (15.3%) | |
| Missing | 786 (20.8%) | 53 (18.1%) | 225 (21.3%) | 93 (21.1%) | 185 (21.1%) | 230 (20.8%) | |

| | | | | | | | |
|---|---|---|---|---|---|---|---|
| Smoking status | | | | | | | |
| Never | 1949 (51.7%) | 65 (22.2%) | 732 (69.3%) | 233 (53.0%) | 446 (50.9%) | 473 (42.8%) | |
| Former smoker | 867 (23.0%) | 94 (32.1%) | 225 (21.3%) | 87 (19.8%) | 224 (25.5%) | 237 (21.5%) | <0.001 |
| <10/day | 365 (9.7%) | 36 (12.3%) | 51 (4.8%) | 44 (10.0%) | 91 (10.4%) | 143 (13.0%) | |
| 10-20/day | 280 (7.4%) | 27 (9.2%) | 27 (2.6%) | 47 (10.7%) | 69 (7.9%) | 110 (10.0%) | |
| ≥20/day | 309 (8.2%) | 71 (24.2%) | 21 (2.0%) | 29 (6.6%) | 47 (5.4%) | 141 (12.8%) | |
| Hypertension status | | | | | | | |
| Normal | 1531 (40.6%) | 97 (33.1%) | 533 (50.5%) | 199 (45.2%) | 307 (35.0%) | 395 (35.8%) | |
| Elevated | 614 (16.3%) | 48 (16.4%) | 153 (14.5%) | 57 (13.0%) | 163 (18.6%) | 193 (17.5%) | |
| Hypertension stage 1 | 845 (22.4%) | 59 (20.1%) | 219 (20.7%) | 98 (22.3%) | 221 (25.2%) | 248 (22.5%) | <0.001 |
| Hypertension stage 2 | 634 (16.8%) | 77 (26.3%) | 117 (11.1%) | 66 (15%) | 153 (17.4%) | 221 (20%) | |
| Hypertension crisis | 32 (0.8%) | 6 (2.0%) | 1 (0.1%) | 0 (0.0%) | 6 (0.7%) | 19 (1.7%) | |
| Missing | 114 (3.0%) | 6 (2.0%) | 33 (3.1%) | 20 (4.5%) | 27 (3.1%) | 28 (2.5%) | |

| | | | | | | | |
|---|---|---|---|---|---|---|---|
| Diabetes status | | | | | | | |
| Non-diabetes | 1986 (52.7%) | 102 (34.8%) | 702 (66.5%) | 254 (57.7%) | 407 (46.4%) | 521 (47.2%) | |
| Prediabetes | 1139 (30.2%) | 99 (33.8%) | 244 (23.1%) | 125 (28.4%) | 310 (35.3%) | 361 (32.7%) | |
| Diabetes | 286 (7.6%) | 37 (12.6%) | 52 (4.9%) | 22 (5.0%) | 71 (8.1%) | 104 (9.4%) | <0.001 |
| Poorly controlled diabetes | 200 (5.3%) | 30 (10.2%) | 22 (2.1%) | 13 (3.0%) | 58 (6.6%) | 77 (7.0%) | |
| Missing | 159 (4.2%) | 25 (8.5%) | 36 (3.4%) | 26 (5.9%) | 31 (3.5%) | 41 (3.7%) | |

*Note*:

Edentulous: individuals with no natural teeth present in oral cavity; Oral health: individuals with neither caries and nor periodontitis, as no periodontitis defined by CDC/AAP definition; Caries: individuals with caries present and with no periodontitis based on CDC/AAP definition; Periodontitis: individuals with no caries and with mild/moderate/severe periodontitis based on CDC/AAP definition; Caries+Periodontitis: individuals with both caries present and with mild/moderate/severe periodontitis based on CDC/AAP definition.

BMI (body mass index): calculated as weight (kg) divided by height squared ($m^2$).

Income-poverty ratio: calculated by dividing total family income by the poverty threshold based on poverty guidelines, specific to family size, year and state.

Hypertension status: categorized based on examined systolic and diastolic blood pressure values (SBP and DBP). Normal: SBP <120 mmHg and DBP <80 mmHg; Elevated: SBP 120-129 mmHg and DBP <80 mmHg; hypertension stage I: SBP 130-139 mmHg or DBP 80-89 mmHg; hypertension stage II: SBP ≥140 mmHg or DBP ≥90 mmHg; hypertension crisis: SBP >180 mmHg and/or DBP >120 mmHg.

Diabetes status: defined by tested glycohemoglobin (HbA1c) levels. Non-diabetes: HbA1c <5.7%; Prediabetes: HbA1c 5.7-6.4%; Diabetes: HbA1c ≥6.5%; Poorly controlled diabetes: HbA1c ≥8.0%.

Table S2. Characteristics of study participants with different oral health conditions based on caries and periodontitis status with ACES periodontitis definitions.

| **ACES definition** | **Overall** | **Edentulous** | **Oral Health** | **Caries** | **Periodontitis** | **Caries+Periodontitis** | ***p* value** |
|---|---|---|---|---|---|---|---|
| **N** | 3770 | 293 | 264 | 118 | 1669 | 1426 | |
| Age | 49 (39-59) | 61 (54-65) | 40 (33-48) | 38 (34-44) | 50 (40-60) | 49 (40-58) | <0.001 |
| Sex | | | | | | | |
| Male | 1955 (51.9%) | 154 (52.6%) | 82 (31.1%) | 39 (33.1%) | 836 (50.1%) | 844 (59.2%) | <0.001 |
| Female | 1815 (48.1%) | 139 (47.4%) | 182 (68.9%) | 79 (66.9%) | 833 (49.9%) | 582 (40.8%) | |

| | | | | | | | |
|---|---|---|---|---|---|---|---|
| BMI (kg/m$^2$) | 28.3 (24.7-32.9) | 27.7 (24.5-33.1) | 26.9 (23.5-31.1) | 29.6 (26.7-33.5) | 28 (24.6-32.5) | 28.8 (25.1-33.5) | <0.001 |
| Income-poverty ratio | 1.8 (1.0-4.0) | 1.1 (0.8-2.0) | 3.7 (2.1-5.0) | 1.3 (0.8-3.1) | 2.6 (1.3-4.9) | 1.3 (0.8-2.6) | <0.001 |
| Education level | | | | | | | |
| Less than 9th grade | 433 (11.5%) | 56 (19.1%) | 6 (2.3%) | 9 (7.6%) | 138 (8.3%) | 224 (15.7%) | |
| 9-11th grade | 632 (16.8%) | 84 (28.7%) | 13 (4.9%) | 19 (16.1%) | 190 (11.4%) | 326 (22.9%) | |
| High school grad/GED or Equivalent | 834 (22.1%) | 73 (24.9%) | 32 (12.1%) | 41 (34.7%) | 321 (19.2%) | 367 (25.7%) | |
| Some college or AA degree | 1013 (26.9%) | 64 (21.8%) | 68 (25.8%) | 39 (33.1%) | 479 (28.7%) | 363 (25.5%) | <0.001 |
| College graduate or above | 856 (22.7%) | 16 (5.5%) | 145 (54.9%) | 10 (8.5%) | 540 (32.4%) | 145 (10.2%) | |
| Missing | 2 (0.1%) | 0 (0.0%) | 0 (0.0%) | 0 (0.0%) | 1 (0.1%) | 1 (0.1%) | |
| Alcohol consumption | | | | | | | <0.001 |
| Never | 687 (18.2%) | 103 (35.2%) | 24 (9.1%) | 23 (19.5%) | 243 (14.6%) | 294 (20.6%) | |

| | | | | | | | |
|---|---|---|---|---|---|---|---|
| Everyday/drink | 158 (4.2%) | 11 (3.8%) | 4 (1.5%) | 3 (2.5%) | 65 (3.9%) | 75 (5.3%) | |
| Every week/drink | 930 (24.7%) | 41 (14.0%) | 92 (34.8%) | 19 (16.1%) | 431 (25.8%) | 347 (24.3%) | |
| Every month/drink | 532 (14.1%) | 28 (9.6%) | 52 (19.7%) | 12 (10.2%) | 257 (15.4%) | 183 (12.8%) | |
| Several month/drink | 677 (18.0%) | 57 (19.5%) | 43 (16.3%) | 36 (30.5%) | 312 (18.7%) | 229 (16.1%) | |
| Missing | 786 (20.8%) | 53 (18.1%) | 49 (18.6%) | 25 (21.2%) | 361 (21.6%) | 298 (20.9%) | |
| Smoking status | | | | | | | <0.001 |
| Never | 1949 (51.7%) | 65 (22.2%) | 191 (72.3%) | 69 (58.5%) | 987 (59.1%) | 637 (44.7%) | |
| Former smoker | 867 (23%) | 94 (32.1%) | 52 (19.7%) | 18 (15.3%) | 397 (23.8%) | 306 (21.5%) | |
| <10/day | 365 (9.7%) | 36 (12.3%) | 11 (4.2%) | 12 (10.2%) | 131 (7.8%) | 175 (12.3%) | |
| 10-20/day | 280 (7.4%) | 27 (9.2%) | 6 (2.3%) | 9 (7.6%) | 90 (5.4%) | 148 (10.4%) | |
| ≥20/day | 309 (8.2%) | 71 (24.2%) | 4 (1.5%) | 10 (8.5%) | 64 (3.8%) | 160 (11.2%) | |
| Hypertension status | | | | | | | <0.001 |
| Normal | 1531 (40.6%) | 97 (33.1%) | 145 (54.9%) | 64 (54.2%) | 695 (41.6%) | 530 (37.2%) | |

| | | | | | | | |
|---|---|---|---|---|---|---|---|
| Elevated | 614 (16.3%) | 48 (16.4%) | 44 (16.7%) | 10 (8.5%) | 272 (16.3%) | 240 (16.8%) | |
| Hypertension stage 1 | 845 (22.4%) | 59 (20.1%) | 44 (16.7%) | 24 (20.3%) | 396 (23.7%) | 322 (22.6%) | |
| Hypertension stage 2 | 634 (16.8%) | 77 (26.3%) | 21 (8%) | 15 (12.7%) | 249 (14.9%) | 272 (19.1%) | |
| Hypertension crisis | 32 (0.8%) | 6 (2%) | 0 (0.0%) | 0 (0.0%) | 7 (0.4%) | 19 (1.3%) | |
| Missing | 114 (3%) | 6 (2%) | 10 (3.8%) | 5 (4.2%) | 50 (3%) | 43 (3%) | |
| Diabetes status | | | | | | | |
| Non-diabetes | 1986 (52.7%) | 102 (34.8%) | 187 (70.8%) | 77 (65.3%) | 922 (55.2%) | 698 (48.9%) | |
| Prediabetes | 1139 (30.2%) | 99 (33.8%) | 60 (22.7%) | 31 (26.3%) | 494 (29.6%) | 455 (31.9%) | |
| Diabetes | 286 (7.6%) | 37 (12.6%) | 8 (3.0%) | 5 (4.2%) | 115 (6.9%) | 121 (8.5%) | <0.001 |
| Poorly controlled diabetes | 200 (5.3%) | 30 (10.2%) | 3 (1.1%) | 0 (0.0%) | 77 (4.6%) | 90 (6.3%) | |
| Missing | 159 (4.2%) | 25 (8.5%) | 6 (2.3%) | 5 (4.2%) | 61 (3.7%) | 62 (4.3%) | |

*Note*:

Edentulous: individuals with no natural teeth present in oral cavity; Oral health: individuals with neither caries and nor periodontitis, as stage I periodontitis were considered as no periodontitis defined by ACES definition; Caries: individuals with caries present and with stage I periodontitis based on ACES definition; Periodontitis: individuals with no caries and with stage II-IV periodontitis based on ACES definition; Caries+Periodontitis: individuals with both caries present and with stage II-IV periodontitis based on ACES definition.

BMI (body mass index): calculated as weight (kg) divided by height squared ($m^2$).

Income-poverty ratio: calculated by dividing total family income by the poverty threshold based on poverty guidelines, specific to family size, year and state.

Hypertension status: categorized based on examined systolic and diastolic blood pressure values (SBP and DBP). Normal: SBP <120 mmHg and DBP <80 mmHg; Elevated: SBP 120-129 mmHg and DBP <80 mmHg; hypertension stage I: SBP 130-139 mmHg or DBP 80-89 mmHg; hypertension stage II: SBP ≥140 mmHg or DBP ≥90 mmHg; hypertension crisis: SBP >180 mmHg and/or DBP >120 mmHg.

Diabetes status: defined by tested glycohemoglobin (HbA1c) levels. Non-diabetes: HbA1c <5.7%; Prediabetes: HbA1c 5.7-6.4%; Diabetes: HbA1c ≥6.5%; Poorly controlled diabetes: HbA1c ≥8.0%.

Table S3. Characteristics of study participants without caries and with different status of periodontitis according to CDC/AAP periodontitis definitions.

| CDC/AAP definition | Overall | No Periodontitis | Mild Periodontitis | Moderate Periodontitis | Severe Periodontitis | *p* value |
|---|---|---|---|---|---|---|
| **N** | 1933 | 1056 | 72 | 592 | 213 | |
| Age | 48 (39-58) | 44 (36-54) | 43 (37-52) | 53 (44-62) | 54 (46-62) | <0.001 |
| Sex | | | | | | |
| Male | 918 (47.5%) | 417 (39.5%) | 43 (59.7%) | 300 (50.7%) | 158 (74.2%) | <0.001 |
| Female | 1015 (52.5%) | 639 (60.5%) | 29 (40.3%) | 292 (49.3%) | 55 (25.8%) | |
| BMI (kg/m$^2$) | 27.8 (24.5-32.3) | 27.3 (24-31.8) | 28.8 (25.9-34.1) | 28.8 (25-33.2) | 27.7 (24.4-31.7) | <0.001 |
| Income-poverty ratio | 2.8 (1.3-5.0) | 3.6 (1.6-5.0) | 2.2 (1.3-4.3) | 2.2 (1.2-4.4) | 1.7 (1.1-3.2) | <0.001 |
| Education level | | | | | | |
| Less than 9th grade | 144 (7.4%) | 46 (4.4%) | 5 (6.9%) | 62 (10.5%) | 31 (14.6%) | |
| 9-11th grade | 203 (10.5%) | 86 (8.1%) | 3 (4.2%) | 83 (14.0%) | 31 (14.6%) | <0.001 |
| High school grad/GED or Equivalent | 353 (18.3%) | 140 (13.3%) | 18 (25.0%) | 136 (23%) | 59 (27.7%) | |

| | | | | | | |
|---|---|---|---|---|---|---|
| Some college or AA degree | 547 (28.3%) | 294 (27.8%) | 23 (31.9%) | 169 (28.5%) | 61 (28.6%) | |
| College graduate or above | 685 (35.4%) | 490 (46.4%) | 23 (31.9%) | 141 (23.8%) | 31 (14.6%) | |
| Missing | 1 (0.1%) | 0 (0.0%) | 0 (0.0%) | 1 (0.2%) | 0 (0.0%) | |
| Alcohol consumption | | | | | <0.001 | |
| Never | 267 (13.8%) | 112 (10.6%) | 6 (8.3%) | 109 (18.4%) | 40 (18.8%) | |
| Everyday/drink | 69 (3.6%) | 25 (2.4%) | 2 (2.8%) | 26 (4.4%) | 16 (7.5%) | |
| Every week/drink | 523 (27.1%) | 309 (29.3%) | 20 (27.8%) | 126 (21.3%) | 68 (31.9%) | |
| Every month/drink | 309 (16.0%) | 192 (18.2%) | 12 (16.7%) | 79 (13.3%) | 26 (12.2%) | |
| Several month/drink | 355 (18.4%) | 193 (18.3%) | 17 (23.6%) | 117 (19.8%) | 28 (13.1%) | <0.001 |
| Missing | 410 (21.2%) | 225 (21.3%) | 15 (20.8%) | 135 (22.8%) | 35 (16.4%) | |
| Smoking status | | | | | <0.001 | |
| Never | 1178 (60.9%) | 732 (69.3%) | 53 (73.6%) | 312 (52.7%) | 81 (38.0%) | <0.001 |

| | | | | | | |
|---|---|---|---|---|---|---|
| Former smoker | 449 (23.2%) | 225 (21.3%) | 10 (13.9%) | 149 (25.2%) | 65 (30.5%) | |
| <10/day | 142 (7.3%) | 51 (4.8%) | 3 (4.2%) | 57 (9.6%) | 31 (14.6%) | |
| 10-20/day | 96 (5.0%) | 27 (2.6%) | 4 (5.6%) | 45 (7.6%) | 20 (9.4%) | |
| ≥20/day | 68 (3.5%) | 21 (2.0%) | 2 (2.8%) | 29 (4.9%) | 16 (7.5%) | |
| Hypertension status | | | | | <0.001 | |
| Normal | 840 (43.5%) | 533 (50.5%) | 30 (41.7%) | 208 (35.1%) | 69 (32.4%) | |
| Elevated | 316 (16.3%) | 153 (14.5%) | 13 (18.1%) | 109 (18.4%) | 41 (19.2%) | |
| Hypertension stage 1 | 440 (22.8%) | 219 (20.7%) | 17 (23.6%) | 159 (26.9%) | 45 (21.1%) | |
| | | | | | | <0.001 |
| Hypertension stage 2 | 270 (14.0%) | 117 (11.1%) | 9 (12.5%) | 94 (15.9%) | 50 (23.5%) | |
| Hypertension crisis | 7 (0.4%) | 1 (0.1%) | 0 (0.0%) | 3 (0.5%) | 3 (1.4%) | |
| Missing | 60 (3.1%) | 33 (3.1%) | 3 (4.2%) | 19 (3.2%) | 5 (2.3%) | |
| Diabetes status | | | | | | |

| | | | | | | |
|---|---|---|---|---|---|---|
| Non-diabetes | 1109 (57.4%) | 702 (66.5%) | 46 (63.9%) | 269 (45.4%) | 92 (43.2%) | |
| Prediabetes | 554 (28.7%) | 244 (23.1%) | 18 (25.0%) | 218 (36.8%) | 74 (34.7%) | |
| Diabetes | 123 (6.4%) | 52 (4.9%) | 5 (6.9%) | 46 (7.8%) | 20 (9.4%) | |
| Poorly controlled diabetes | 80 (4.1%) | 22 (2.1%) | 2 (2.8%) | 36 (6.1%) | 20 (9.4%) | <0.001 |
| Missing | 67 (3.5%) | 36 (3.4%) | 1 (1.4%) | 23 (3.9%) | 7 (3.3%) | |

*Note*:

Under the CDC/AAP definition, no periodontitis: individual absence of mild, moderate, or severe periodontitis found; Mild periodontitis: individual with ≥2 interproximal sites with attachment loss ≥3 mm, and ≥2 interproximal sites with pocket depth ≥4 mm (not on same tooth) or one site with pocket depth ≥5 mm; Moderate periodontitis: individual with ≥2 interproximal sites with attachment loss ≥4 mm (not on same tooth), or ≥2 interproximal sites with pocket depth ≥5 mm (not on same tooth); Severe periodontitis: individual with ≥2 interproximal sites with attachment loss ≥6 mm (not on same tooth) and ≥1 interproximal site with pocket depth ≥5 mm.

BMI (body mass index): calculated as weight (kg) divided by height squared ($m^2$).

Income-poverty ratio: calculated by dividing total family income by the poverty threshold based on poverty guidelines, specific to family size, year and state.

Hypertension status: categorized based on examined systolic and diastolic blood pressure values (SBP and DBP). Normal: SBP <120 mmHg and DBP <80 mmHg; Elevated: SBP 120-129 mmHg and DBP <80 mmHg; hypertension stage I: SBP 130-139 mmHg or DBP 80-89 mmHg; hypertension stage II: SBP ≥140 mmHg or DBP ≥90 mmHg; hypertension crisis: SBP >180 mmHg and/or DBP >120 mmHg.

Diabetes status: defined by tested glycohemoglobin (HbA1c) levels. Non-diabetes: HbA1c <5.7%; Prediabetes: HbA1c 5.7-6.4%; Diabetes: HbA1c ≥6.5%; Poorly controlled diabetes: HbA1c ≥8.0%.

Table S4. Characteristics of study participants without caries and with different status of periodontitis according to ACES periodontitis definitions.

| **ACES definition** | **Overall** | **Stage I Periodontitis** | **Stage II Periodontitis** | **Stage III/IV Periodontitis** | ***p* value** |
|---|---|---|---|---|---|
| **N** | 1933 | 264 | 1000 | 669 | |
| Age | 48 (39-58) | 40 (33-48) | 46 (37-56) | 55 (45-62) | <0.001 |
| Sex | | | | | |
| Male | 918 (47.5%) | 82 (31.1%) | 445 (44.5%) | 391 (58.4%) | <0.001 |
| Female | 1015 (52.5%) | 182 (68.9%) | 555 (55.5%) | 278 (41.6%) | |

| | | | | | |
|---|---|---|---|---|---|
| BMI (kg/m$^2$) | 27.8 (24.5-32.3) | 26.9 (23.5-31.1) | 27.8 (24.5-32.1) | 28.2 (25.0-32.8) | 0.003 |
| Income-poverty ratio | 2.8 (1.3-5.0) | 3.7 (2.1-5.0) | 3.1 (1.4-5.0) | 2.0 (1.1-4.0) | <0.001 |
| Education level | | | | | <0.001 |
| Less than 9th grade | 144 (7.4%) | 6 (2.3%) | 57 (5.7%) | 81 (12.1%) | |
| 9-11th grade | 203 (10.5%) | 13 (4.9%) | 92 (9.2%) | 98 (14.6%) | |
| High school grad/GED or Equivalent | 353 (18.3%) | 32 (12.1%) | 146 (14.6%) | 175 (26.2%) | |
| Some college or AA degree | 547 (28.3%) | 68 (25.8%) | 295 (29.5%) | 184 (27.5%) | |
| College graduate or above | 685 (35.4%) | 145 (54.9%) | 410 (41.0%) | 130 (19.4%) | |
| Missing | 1 (0.1%) | 0 (0.0%) | 0 (0.0%) | 1 (0.1%) | |
| Alcohol consumption | | | | | <0.001 |
| Never | 267 (13.8%) | 24 (9.1%) | 115 (11.5%) | 128 (19.1%) | |

| | | | | | |
|---|---|---|---|---|---|
| Everyday/drink | 69 (3.6%) | 4 (1.5%) | 28 (2.8%) | 37 (5.5%) | |
| Every week/drink | 523 (27.1%) | 92 (34.8%) | 264 (26.4%) | 167 (25.0%) | |
| Every month/drink | 309 (16.0%) | 52 (19.7%) | 174 (17.4%) | 83 (12.4%) | |
| Several month/drink | 355 (18.4%) | 43 (16.3%) | 187 (18.7%) | 125 (18.7%) | |
| Missing | 410 (21.2%) | 49 (18.6%) | 232 (23.2%) | 129 (19.3%) | |
| Smoking status | | | | | |
| Never | 1178 (60.9%) | 191 (72.3%) | 673 (67.3%) | 314 (46.9%) | |
| Former smoker | 449 (23.2%) | 52 (19.7%) | 219 (21.9%) | 178 (26.6%) | <0.001 |
| <10/day | 142 (7.3%) | 11 (4.2%) | 54 (5.4%) | 77 (11.5%) | |
| 10-20/day | 96 (5.0%) | 6 (2.3%) | 33 (3.3%) | 57 (8.5%) | |
| ≥20/day | 68 (3.5%) | 4 (1.5%) | 21 (2.1%) | 43 (6.4%) | |
| Hypertension status | | | | | |
| Normal | 840 (43.5%) | 145 (54.9%) | 476 (47.6%) | 219 (32.7%) | <0.001 |

| | | | | | |
|---|---|---|---|---|---|
| Elevated | 316 (16.3%) | 44 (16.7%) | 139 (13.9%) | 133 (19.9%) | |
| Hypertension stage 1 | 440 (22.8%) | 44 (16.7%) | 227 (22.7%) | 169 (25.3%) | |
| Hypertension stage 2 | 270 (14.0%) | 21 (8.0%) | 123 (12.3%) | 126 (18.8%) | |
| Hypertension crisis | 7 (0.4%) | 0 (0.0%) | 1 (0.1%) | 6 (0.9%) | |
| Missing | 60 (3.1%) | 10 (3.8%) | 34 (3.4%) | 16 (2.4%) | |
| Diabetes status | | | | | |
| Non-diabetes | 1109 (57.4%) | 187 (70.8%) | 625 (62.5%) | 297 (44.4%) | |
| Prediabetes | 554 (28.7%) | 60 (22.7%) | 258 (25.8%) | 236 (35.3%) | |
| Diabetes | 123 (6.4%) | 8 (3.0%) | 56 (5.6%) | 59 (8.8%) | |
| Poorly controlled diabetes | 80 (4.1%) | 3 (1.1%) | 25 (2.5%) | 52 (7.8%) | <0.001 |
| Missing | 67 (3.5%) | 6 (2.3%) | 36 (3.6%) | 25 (3.7%) | |

*Note*:

Under the ACES definition, stage I periodontitis: individual with interproximal attachment loss ≥1 mm at ≥2 non-adjacent teeth, with maximum attachment loss of 1-2 mm; Stage II periodontitis: individual with interproximal attachment loss ≥1 mm at ≥2 non-adjacent teeth, with maximum attachment loss of 3-4 mm; Stage III/IV periodontitis: individual with interproximal attachment loss ≥1 mm at ≥2 non-adjacent teeth, with maximum attachment loss ≥5 mm.

BMI (body mass index): calculated as weight (kg) divided by height squared ($m^2$).

Income-poverty ratio: calculated by dividing total family income by the poverty threshold based on poverty guidelines, specific to family size, year and state.

Hypertension status: categorized based on examined systolic and diastolic blood pressure values (SBP and DBP). Normal: SBP <120 mmHg and DBP <80 mmHg; Elevated: SBP 120-129 mmHg and DBP <80 mmHg; hypertension stage I: SBP 130-139 mmHg or DBP 80-89 mmHg; hypertension stage II: SBP ≥140 mmHg or DBP ≥90 mmHg; hypertension crisis: SBP >180 mmHg and/or DBP >120 mmHg.

Diabetes status: defined by tested glycohemoglobin (HbA1c) levels. Non-diabetes: HbA1c <5.7%; Prediabetes: HbA1c 5.7-6.4%; Diabetes: HbA1c ≥6.5%; Poorly controlled diabetes: HbA1c ≥8.0%.

**See in attached files:**

Supplementary Spreadsheet 1-differential abundance by DESeq2 in different oral conditions

Supplementary Spreadsheet 2-differential abundance by ALDEx2 in different oral conditions

Supplementary Spreadsheet 3-differential abundance by DESeq2 in different periodontitis severity.

Supplementary Spreadsheet 4-differential abundance by ALDEx2 in different periodontitis severity.
